# Caregiving transition trajectories and health behaviours: a latent class analysis of the UK Household Longitudinal Study

**DOI:** 10.64898/2026.09.26.26364079

**Authors:** Enrico Pfeifer, Rebecca E Lacey, Baowen Xue, Hynek Pikhart, Anne McMunn

## Abstract

**Background:** Unpaid caregiving is a recognised social determinant of health, yet most research treats it as a fixed exposure measured at a single time point, and little is known about how patterns of caregiving transitions relate to health behaviours.

**Methods:** Secondary analysis of the UK Household Longitudinal Study (waves 2–13, approximately 2010–2023; n=25,049). Latent class analysis was applied to binary caregiving status across 12 waves to identify caregiving trajectories. Associations between trajectories and four health behaviours (physical inactivity, fruit and vegetable consumption, problematic drinking, current smoking) at waves 7 to 13 were estimated using logistic and linear regression, adjusting for sociodemographic, health and baseline behavioural confounders.

**Results:** An eight-class solution identified a non-caregiving class comprising 62.9% (n=15,760) of the sample. Among caregivers (n=9,289), trajectories were Temporary (20.0%), Former-short (20.2%), Former-long (10.9%), Emerging-short (15.4%), Emerging- long (11.6%), Long-term (15.7%) and Recurrent (6.3%). In fully adjusted models, Recurrent caregivers had higher odds of smoking (OR=1.67, 95%CI: 1.17 to 2.40) compared with non- caregivers but lower odds of physical inactivity (OR=0.65, 95%CI: 0.53 to 0.81) and problematic drinking (OR=0.75, 95%CI: 0.59 to 0.94). Associations for other trajectory classes were largely attenuated after adjustment.

**Conclusion:** The pattern and timing of caregiving transitions are an under-explored dimension of the caregiving experience. Recurrent caregivers had a distinctive profile, with elevated smoking but lower physical inactivity and problematic drinking. Recurrent caregiving should be recognised alongside intensity and duration, and support systems better equipped for people who move into and out of care repeatedly.

**What is already known on this topic:**

- Unpaid caregiving has been linked to adverse health outcomes, but most studies treat caregiving as a fixed, single-time-point exposure and rely on researcher-defined categories.
- Evidence on the association between caregiving and health behaviours is inconsistent and largely cross-sectional. The dynamic nature of caregiving, including recurrent episodes, has not been examined in relation to health behaviours.

**What this study adds:**

- Latent class analysis of 12 waves of panel data identified eight distinct caregiving trajectories, capturing variation in the timing, duration, and recurrence of caregiving episodes.
- Recurrent caregivers had higher odds of smoking but lower odds of physical inactivity and problematic drinking, a profile not identifiable through conventional classification approaches.

**How this study might affect research, practice or policy:**

- Recurrent caregiving should be recognised as a distinct dimension of the caregiving experience, and support systems should prioritise reducing the burden of care for people who move into and out of caregiving repeatedly.
- Targeted smoking cessation support may complement upstream support measures but should not replace them.

## Introduction

Unpaid caregiving represents a substantial and growing public health challenge across high-income countries. In the United Kingdom, an estimated 6.5 million adults provide unpaid care to family members, friends, or neighbours with long-term illness, disability, or age-related needs.^1^ Caregiving is increasingly recognised as a social determinant of health, with implications for physical and mental wellbeing that extend across the lifecourse.^2^

Intensive Caregiving has been linked to elevated psychological distress and poorer self-rated health ^3,4^ but the mechanisms through which it affects health remain unclear. Health behaviours represent a plausible pathway. The stress process model posits that the subjective and objective burdens of care may trigger maladaptive coping responses such as increased smoking or alcohol consumption,^5^ while the COM-B model implies that caregiving may erode the capability, opportunity, and motivation required for health-promoting behaviour.^6^ From a lifecourse perspective, transitions into and out of social roles are critical timepoints at which established routines may be disrupted or reshaped.^7^ Collectively, these perspectives imply that changes in caregiving status, and particularly repeated transitions, may influence physical activity, diet, alcohol consumption, and smoking.

Despite this theoretical plausibility, longitudinal evidence remains scarce and inconsistent. Physical activity was higher among caregivers in US and European studies^8,9^ but lower among high-intensity caregivers in Japan,^10^ while entering caregiving was linked to increased problematic drinking^9,10^ and to either no change^10^ or a decrease in smoking.^9^ All of these studies examined entry into caregiving in adults over the age of 50. The only longitudinal study to assess both entry and exit caregiving across the adult lifecourse found that entering caregiving was associated with lower odds of physical inactivity and higher odds of smoking, whereas exiting was associated with a return to higher inactivity.^11^ That study used fixed effects models centred on the first observed transition, however, and so captured a single entry or exit rather than the recurrent and prolonged episodes that characterise caregiving over time.

A more fundamental limitation of this body of work is its treatment of caregiving as a fixed exposure. UK longitudinal data indicate that roughly 7% of adults enter caregiving and around 6% exit in any given year, underlining how frequently these roles change.^12^Even where longer patterns are considered, definitions such as long-term caregiving rely on arbitrary cut-offs that vary across studies, with no agreed thresholds and, hence, limited comparability. Moreover, existing longitudinal studies have typically examined single transitions, such as entry or exit, rather than modelling entire caregiving trajectories..^13^ Therefore, it is useful to conceptualise caregiving through trajectories than a single event.^7^ Trajectories vary in onset, duration, continuity, and recurrence, and these features may shape engagement in health-promoting and health-adverse behaviours more than caregiving status alone.^14^

To address these gaps, this study had three aims. The first was to identify distinct typologies of unpaid caregiving trajectories. The second was to examine their associations with four health behaviours (physical inactivity, fruit and vegetable consumption, problematic drinking, and current smoking). The third was to test whether these association differed by sex or age group.

We expected several distinct trajectory classes to emerge, including emerging, former, recurrent, and long-term caregiving, with the recurrent and long-term classes representing the greatest cumulative exposure (H1). We expected less favourable behaviours in these two classes, in particular higher odds of smoking and physical inactivity, reflecting disrupted routines and stress-related coping (H2), except for problematic drinking, where lower odds were expected as intensive caregiving demands leave less opportunity for drinking (H3). We further expected stronger association between caregiving and less favourable associations among women, given the gendered division of unpaid care (H4), and in younger age group, for whom caregiving is less socially normative (H5).^4,15–17^

## Methods

### Study design and data source

This study was a secondary analysis of the UK Household Longitudinal Study (UKHLS), also known as Understanding Society.^18^ UKHLS is a nationally representative household panel established in 2009 with around 40,000 households, in which all members aged 16 and over are interviewed annually. Data were drawn from waves 2 to 13 (fieldwork approximately 2010 to 2022), providing up to 12 observations per individual. Wave 1 was excluded because the self-completion questionnaire carrying the health behaviour modules are available only from wave 2 onwards (Supplement 13). The study was conducted and reported in accordance with STROBE guidelines.^19^

### Study population

The analytical sample comprised adults aged 16 and over whose caregiving status was observed at a minimum of four waves, a threshold set to allow entry, exit, and recurrence to be identified. Individuals were additionally required to have a valid baseline measure of the relevant health behaviour at wave 2, or at wave 5 where wave 2 was unavailable, and a corresponding outcome measure at wave 7, 9, 11, or 13. The physical activity, diet, and alcohol modules were fielded only at waves 2, 5, 7, 9, 11, and 13, and a change in question format from wave 7 onwards precluded full harmonisation with the earlier waves.17 Waves 2 and 5 therefore provided the baseline measures and waves 7, 9, 11, and 13 the outcomes.

Wave 12 was excluded because the physical activity and alcohol modules were added mid-fieldwork in response to the COVID-19 pandemic. These criteria yielded an analytical sample of 25,049 individuals; a flowchart is provided in the Supplement 4.

### Exposure: caregiving trajectories

Caregiving status was measured at each wave using two questions, asking whether the respondent looked after or gave special help to anyone living with them who was sick, disabled or elderly, and to someone outside the household. Respondents answering affirmatively to either question were coded as caregivers and those answering no to both as non-caregivers, giving a binary indicator at each of the 12 waves from waves 2-13. Caregiving trajectory class, derived from these 12 indicators using latent class analysis, was the exposure of interest in all models.

### Outcomes

Four health behaviours were examined, each measured at waves 7, 9, 11, or 13, using the latest available valid observation.

Physical inactivity was derived from the short form of the International Physical Activity Questionnaire (IPAQ),^20^ using thresholds consistent with UK Chief Medical Officer guidelines.^21^ Participants reported the frequency and duration of moderate and vigorous activity and were classified as active if they reported at least 75 minutes of vigorous activity per week, 150 minutes of moderate activity, or a combination totalling 150 minutes, and as inactive otherwise.

Dietary behaviour was measured as average daily fruit and vegetable intake. Participants reported the days per week on which they consumed fruit and the portions eaten on those days, with equivalent questions for vegetables, from which a daily average was derived.

Problematic drinking was assessed using the Alcohol Use Disorders Identification Test-Consumption (AUDIT-C), a validated three-item screen scored from 0 to 12 measuring the frequency of drinking, the typical number of drinks per occasion and the frequency of binge drinking.^22^ Participants scoring 3 or more (women) or 4 or more (men) were classified as problematic drinkers, consistent with cut-offs used in previous research.^23,24^

Smoking status was self-reported, with those reporting that they currently smoked classified as current smokers.

### Confounders

Models were adjusted for potential confounders identified from the existing literature and a directed acyclic graph (Supplement 2). Sociodemographic confounders were sex, age group, highest educational qualification, ethnicity, occupational class (National Statistics Socio-economic Classification), equivalised household income quintile, employment status, household size, number of children under age 16? in the household, and cohabiting status. Age was grouped into four lifecourse stages, 16 to 29 (early adulthood), 30 to 49 (early mid-adulthood), 50 to 64 (late mid-adulthood), and 65 and over (late adulthood). Health covariates were self-rated health, psychological distress (General Health Questionnaire-12), and, for the physical inactivity model, the SF-12 physical component summary. All confounders were measured at each participant’s baseline wave, defined as the first wave in which caregiving status was observed.

Adjusted models additionally included a baseline measure of the relevant health behaviour from wave 2 or 5. Because the full outcome instruments were not fielded at these waves, the closest available measure was used, namely walking frequency for physical inactivity, a categorical fruit and vegetable measure for diet, frequency of alcohol consumption for problematic drinking, and self-reported smoking for smoking. Each predicted its corresponding outcome at follow-up. The wave of outcome observation was also included to account for period effects. A detailed variable definition can be found in Supplement 1.

### Statistical analysis

Analysis proceeded in two steps. First, latent class analysis (LCA) was applied to the 12 binary caregiving indicators to identify subgroups sharing similar trajectories (Collins and Lanza, 2009).^25^ Models were estimated for two through 12 classes using the poLCA package in R,^26^ and model selection was informed by fit indices (BIC, AIC, entropy), inspection of an elbow plot, and theoretical interpretability. The selected classes were characterised using sequence index plots, state distribution plots, and modal state plots.

Second, class membership was regressed on the health behaviour outcomes in three models per outcome, unadjusted, adjusted for baseline behaviour only, and adjusted for selected confounders. Logistic regression was used for binary outcomes and linear regression for fruit and vegetable consumption. Interactions between class membership and sex, and between class membership and age group, were tested using Wald tests, with stratified results produced where interactions were statistically significant.

To reduce bias, the complex survey design of UKHLS was accounted for using survey weights, with standard errors adjusted for stratification and clustering. Missing covariate and outcome data were handled by multiple imputation using chained equations (m=10).

Two sensitivity analyses were conducted. First, sequence analysis with clustering was used as an alternative method of classifying trajectories. Second, a rule-based observed-transitions variable, classifying participants by their number of transitions between caregiving and non-caregiving states, served as a simpler comparator. Results of sensitivity analyses are reported in Supplement 10 and 11, and all statistical code is available at *[insert github link after acceptance]*.

## Results

### Latent class model selection

We fitted latent class models with one to twelve classes (Table 1). Model fit improved steadily as classes were added without reaching a clear optimum, with the largest gains up to four classes and an elbow at four or five classes (Supplement 5). Because these solutions did not capture the patterns of interest, higher solutions were compared against our research question and conceptual framework (Supplement 3). A recurrent pattern appeared only in the eight-class solution, whereas the nine-class solution added a class resembling an existing one. We therefore selected the eight-class solution (entropy 0.74; posterior probabilities in Supplement 6).

**Table 1:**
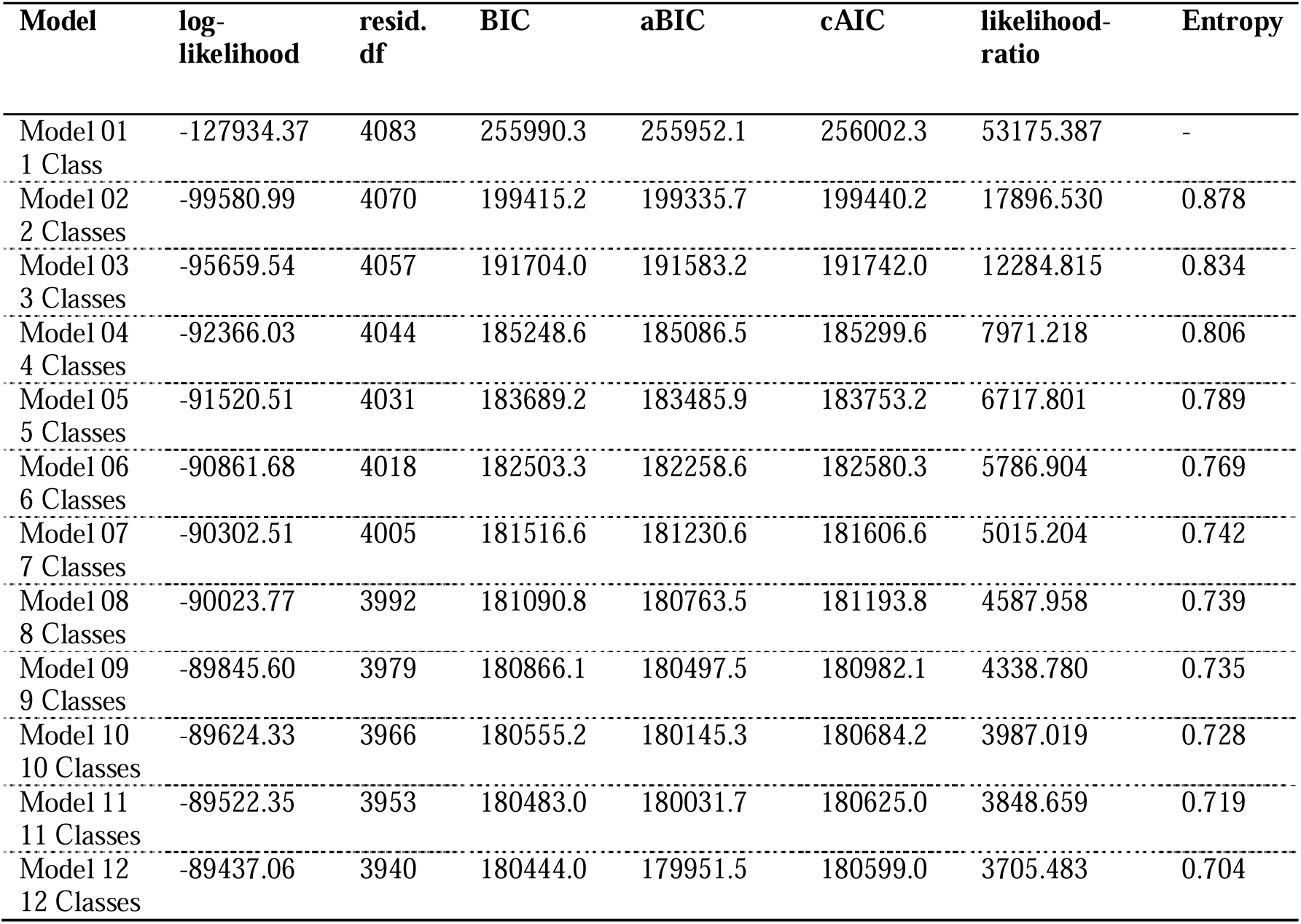
**Latent class model fit statistics** for caregiving status across UKHLS waves 2 to 13 (n=25,049). Models were compared using log-likelihood, Bayesian Information Criterion (BIC), adjusted BIC (aBIC), consistent Akaike Information Criterion (cAIC), likelihood ratio tests, and entropy values.

Classes were labelled on the basis of their posterior probabilities and graphical inspection (Figure 1) as temporary, former-short, former-long, emerging-short, emerging-long, long-term, and recurrent caregivers, alongside a non-caregiving reference class.

**Figure 1:**
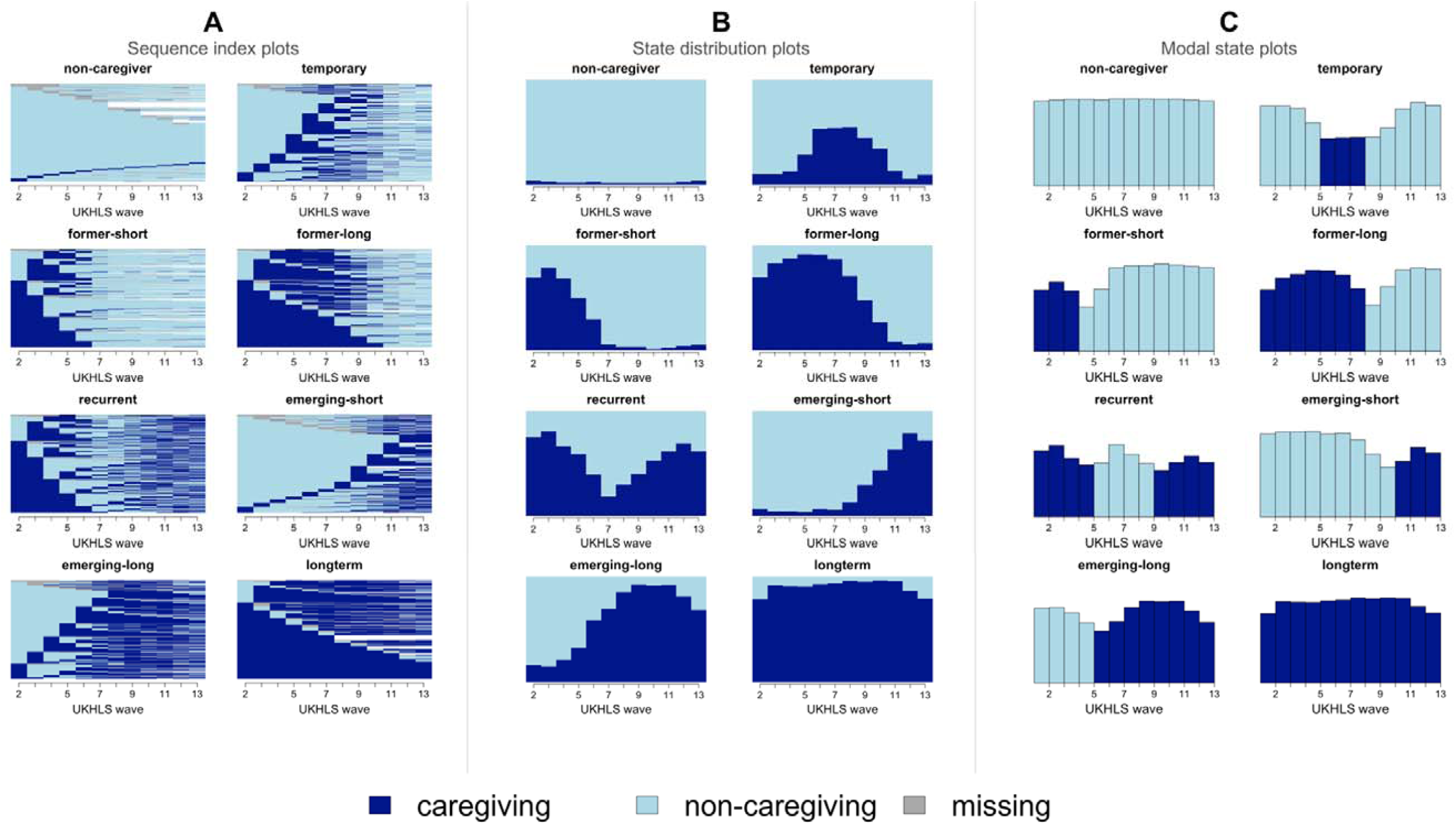
**Caregiving trajectories for the eight-class solution** across UKHLS waves 2 to 13 (n=25,049). Each set of panels represents one latent class, shown through three complementary visualisations of the same classification. (A) Sequence index plot: each horizontal line is an individual participant’s caregiving status trajectory, sorted from the start of the observation window. (B) State distribution plot: the proportion of participants in each caregiving status at every wave. (C) Modal state plot: the most frequent caregiving status at each wave within the class. Dark blue denotes caregiving and light blue non-caregiving. Missing states, shown in grey, appear in the sequence index plot only; the distribution and modal state plots are computed on observed states.

### Descriptive statistics

The analytical sample comprised 25,049 people, with a mean baseline age of 46.5 years; 53.2% were women and 93.6% were white (Table 2). The non-caregiver class was the largest, comprising 62.9% of the sample. Among those classified into a caregiving trajectory, the temporary and former-short classes were the most prevalent (approximately 20% of caregivers each) and the recurrent class the smallest (6.3%). Women were over-represented in every caregiving class relative to non-caregivers, and long-term and former-long caregivers were older, and emerging-short caregivers younger, than the sample as a whole.

**Table 2:**
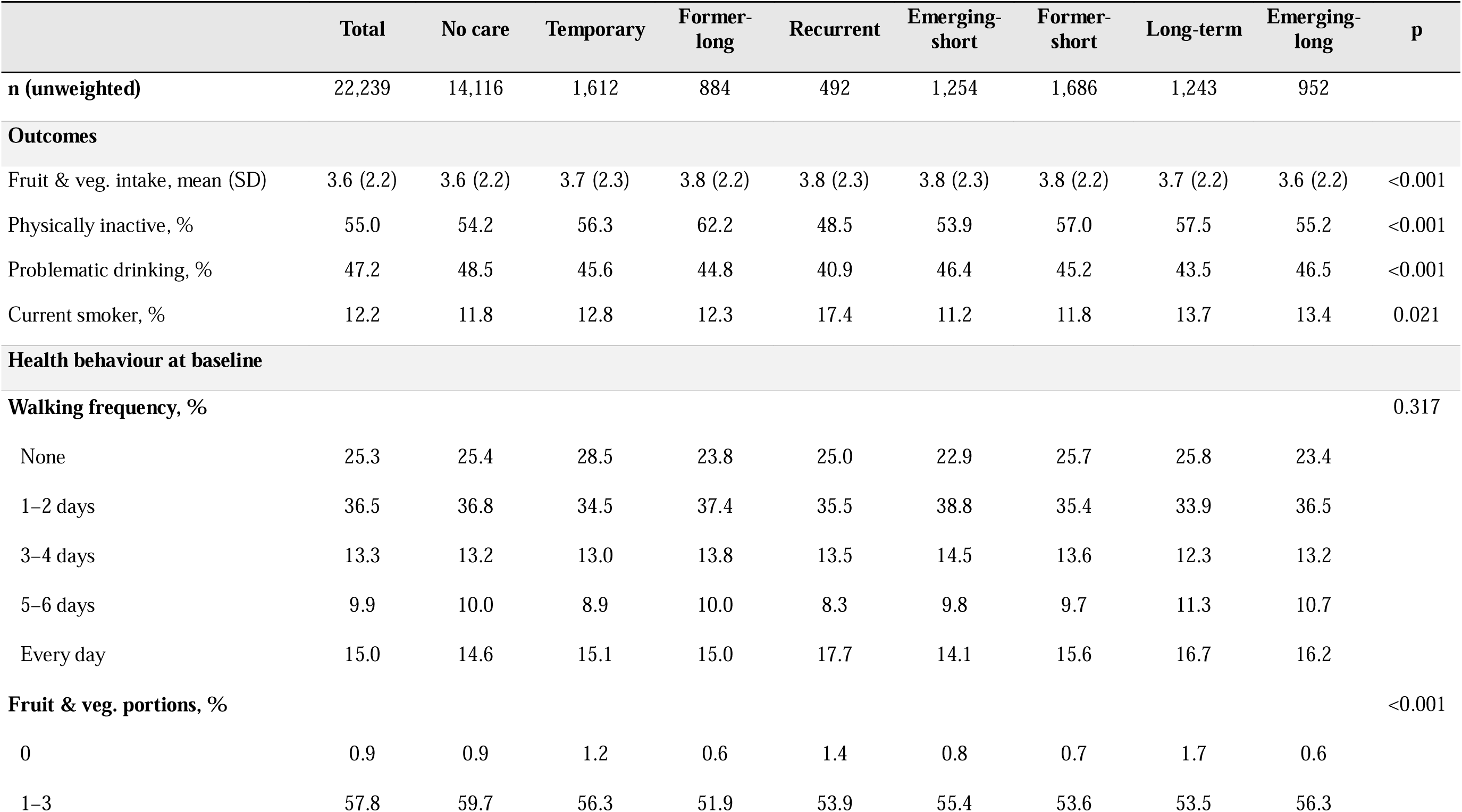
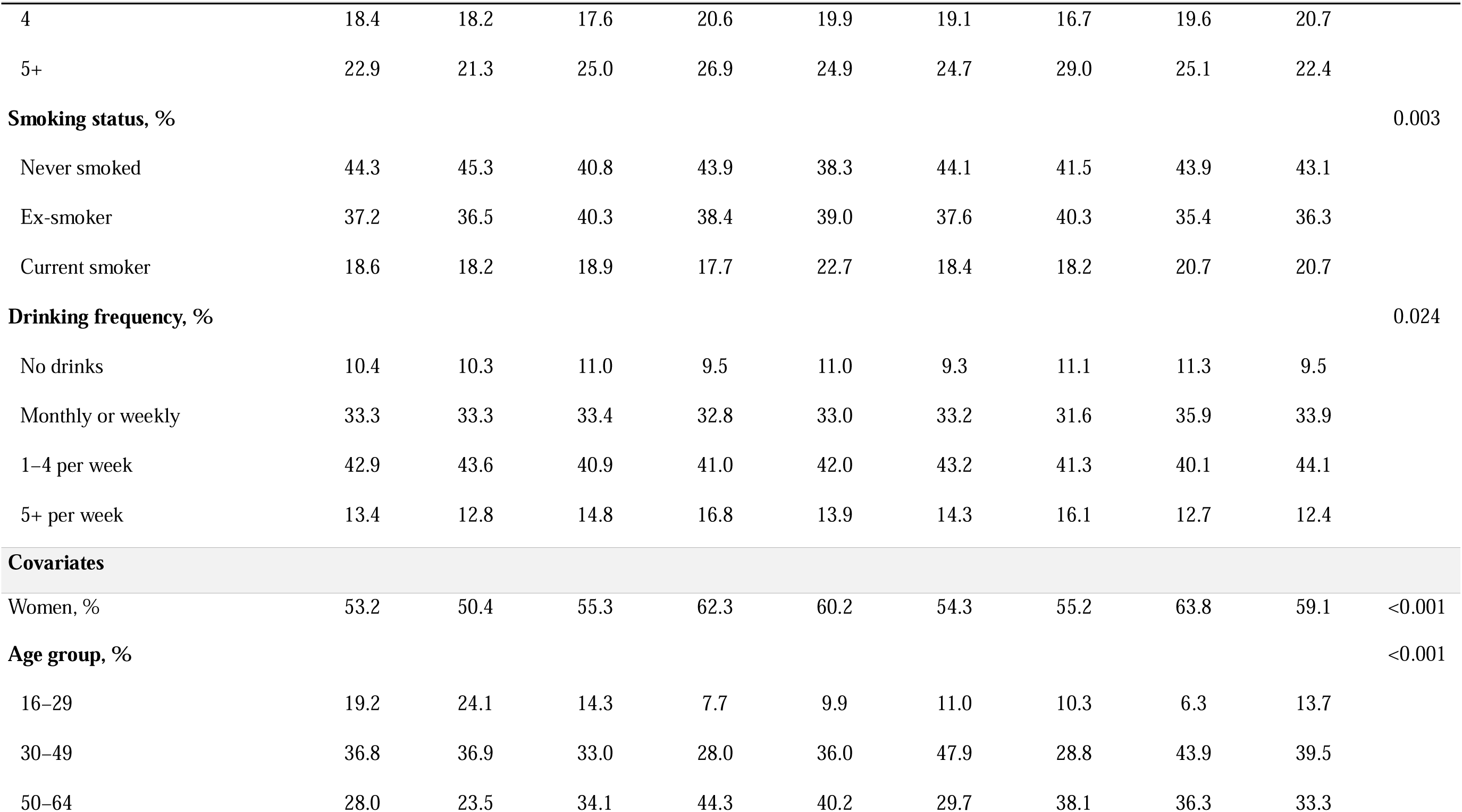
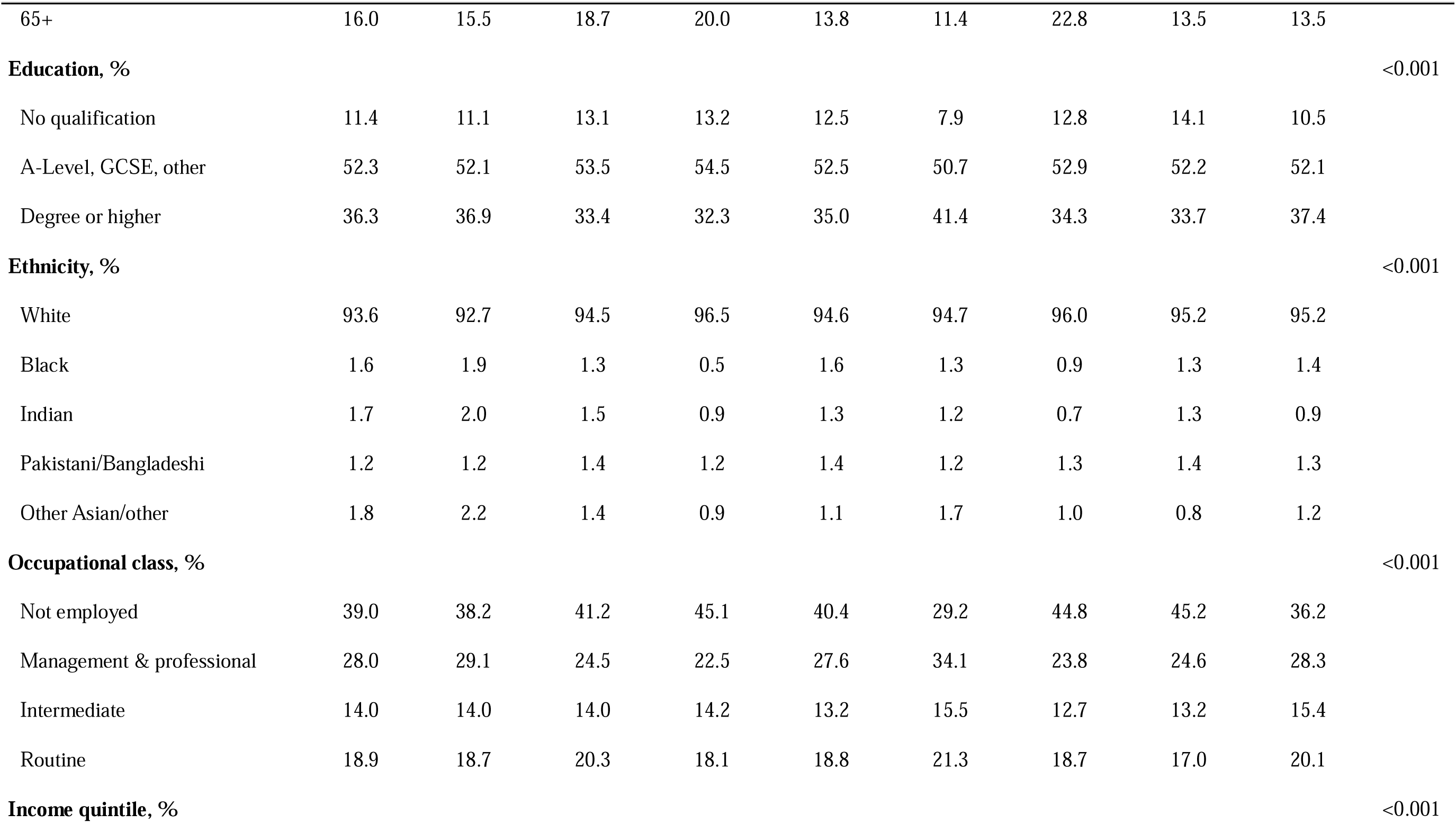
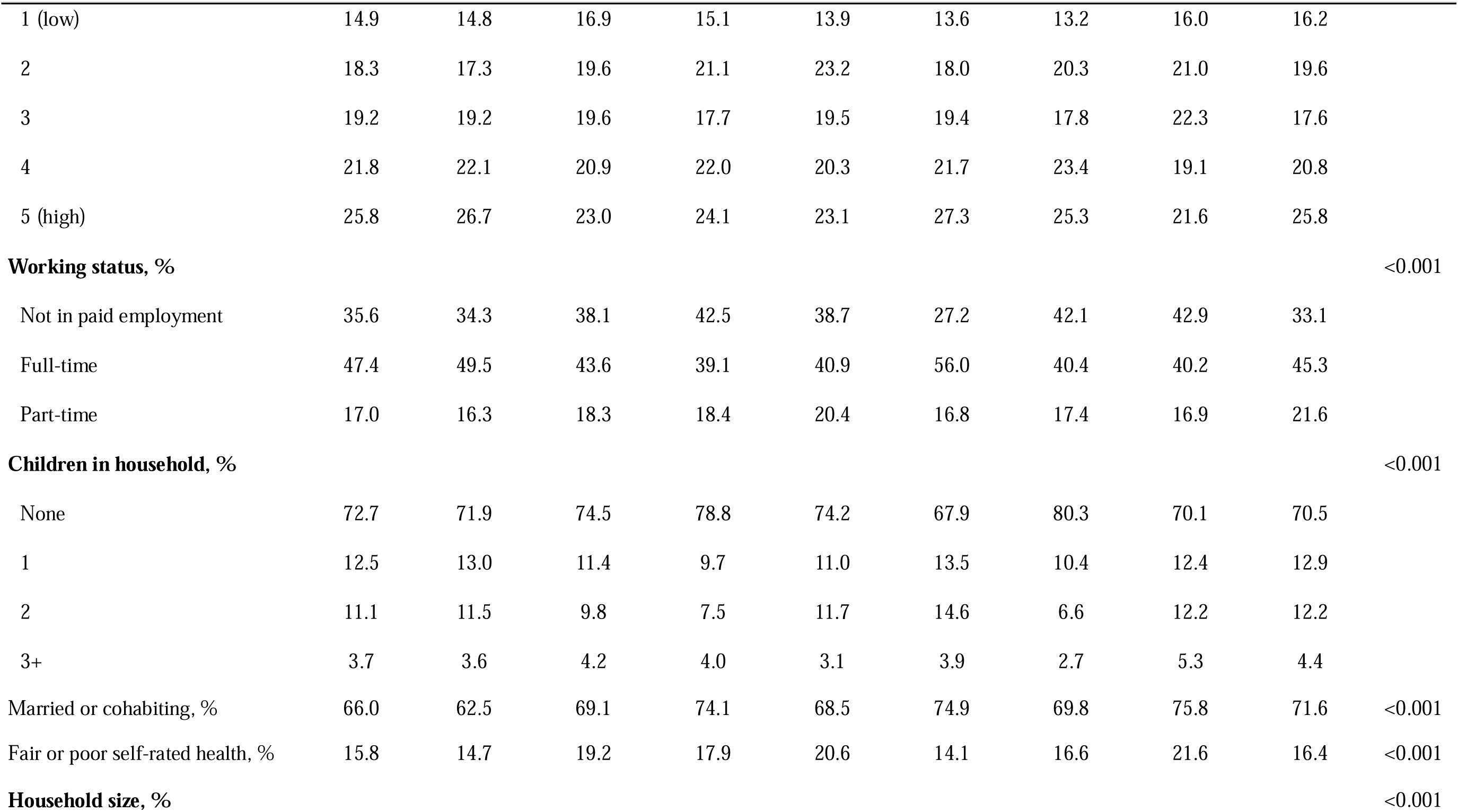
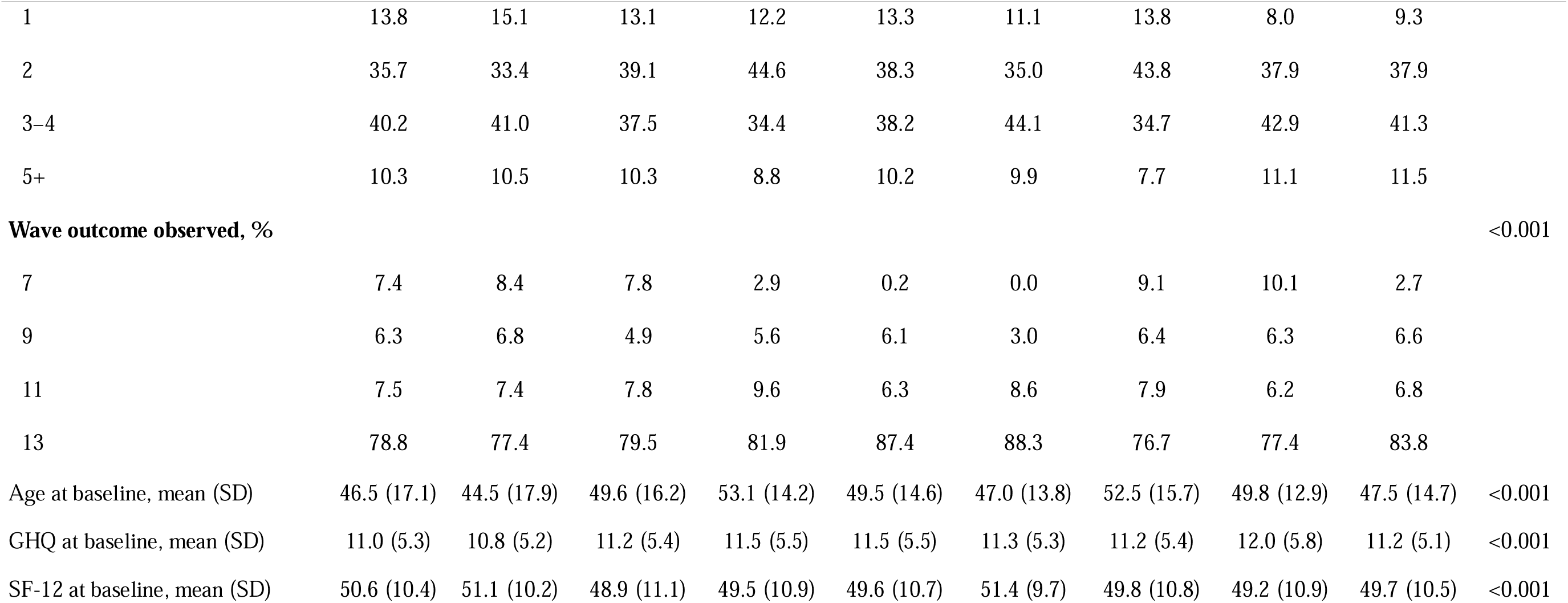
Descriptive statistics for latent classes identified through latent class analysis of caregiving status trajectories (n=25,049), based on pooled results from multiple utation (m=10). Estimates account for complex survey design and clustering at the household level.

|  | Total | No care | Temporary | Former-long | Recurrent | Emerging-short | Former-short | Long-term | Emerging-long | p |
| --- | --- | --- | --- | --- | --- | --- | --- | --- | --- | --- |
| **n (unweighted)** | 22,239 | 14,116 | 1,612 | 884 | 492 | 1,254 | 1,686 | 1,243 | 952 |  |
| **Outcomes** |  |  |  |  |  |  |  |  |  |  |
| Fruit & veg. intake, mean (SD) | 3.6 (2.2) | 3.6 (2.2) | 3.7 (2.3) | 3.8 (2.2) | 3.8 (2.3) | 3.8 (2.3) | 3.8 (2.2) | 3.7 (2.2) | 3.6 (2.2) | <0.001 |
| Physically inactive, % | 55.0 | 54.2 | 56.3 | 62.2 | 48.5 | 53.9 | 57.0 | 57.5 | 55.2 | <0.001 |
| Problematic drinking, % | 47.2 | 48.5 | 45.6 | 44.8 | 40.9 | 46.4 | 45.2 | 43.5 | 46.5 | <0.001 |
| Current smoker, % | 12.2 | 11.8 | 12.8 | 12.3 | 17.4 | 11.2 | 11.8 | 13.7 | 13.4 | 0.021 |
| **Health behaviour at baseline** |  |  |  |  |  |  |  |  |  |  |
| **Walking frequency, %** |  |  |  |  |  |  |  |  |  | 0.317 |
| None | 25.3 | 25.4 | 28.5 | 23.8 | 25.0 | 22.9 | 25.7 | 25.8 | 23.4 |  |
| 1–2 days | 36.5 | 36.8 | 34.5 | 37.4 | 35.5 | 38.8 | 35.4 | 33.9 | 36.5 |  |
| 3–4 days | 13.3 | 13.2 | 13.0 | 13.8 | 13.5 | 14.5 | 13.6 | 12.3 | 13.2 |  |
| 5–6 days | 9.9 | 10.0 | 8.9 | 10.0 | 8.3 | 9.8 | 9.7 | 11.3 | 10.7 |  |
| Every day | 15.0 | 14.6 | 15.1 | 15.0 | 17.7 | 14.1 | 15.6 | 16.7 | 16.2 |  |
| **Fruit & veg. portions, %** |  |  |  |  |  |  |  |  |  | <0.001 |
| 0 | 0.9 | 0.9 | 1.2 | 0.6 | 1.4 | 0.8 | 0.7 | 1.7 | 0.6 |  |
| 1–3 | 57.8 | 59.7 | 56.3 | 51.9 | 53.9 | 55.4 | 53.6 | 53.5 | 56.3 |  |

|  | Total | No care | Temporary | Former-long | Recurrent | Emerging-short | Former-short | Long-term | Emerging-long | p |
| --- | --- | --- | --- | --- | --- | --- | --- | --- | --- | --- |
| 4 | 18.4 | 18.2 | 17.6 | 20.6 | 19.9 | 19.1 | 16.7 | 19.6 | 20.7 |  |
| 5+ | 22.9 | 21.3 | 25.0 | 26.9 | 24.9 | 24.7 | 29.0 | 25.1 | 22.4 |  |
| **Smoking status, %** |  |  |  |  |  |  |  |  |  | 0.003 |
| Never smoked | 44.3 | 45.3 | 40.8 | 43.9 | 38.3 | 44.1 | 41.5 | 43.9 | 43.1 |  |
| Ex-smoker | 37.2 | 36.5 | 40.3 | 38.4 | 39.0 | 37.6 | 40.3 | 35.4 | 36.3 |  |
| Current smoker | 18.6 | 18.2 | 18.9 | 17.7 | 22.7 | 18.4 | 18.2 | 20.7 | 20.7 |  |
| **Drinking frequency, %** |  |  |  |  |  |  |  |  |  | 0.024 |
| No drinks | 10.4 | 10.3 | 11.0 | 9.5 | 11.0 | 9.3 | 11.1 | 11.3 | 9.5 |  |
| Monthly or weekly | 33.3 | 33.3 | 33.4 | 32.8 | 33.0 | 33.2 | 31.6 | 35.9 | 33.9 |  |
| 1–4 per week | 42.9 | 43.6 | 40.9 | 41.0 | 42.0 | 43.2 | 41.3 | 40.1 | 44.1 |  |
| 5+ per week | 13.4 | 12.8 | 14.8 | 16.8 | 13.9 | 14.3 | 16.1 | 12.7 | 12.4 |  |
| **Covariates** |  |  |  |  |  |  |  |  |  |  |
| Women, % | 53.2 | 50.4 | 55.3 | 62.3 | 60.2 | 54.3 | 55.2 | 63.8 | 59.1 | <0.001 |
| **Age group, %** |  |  |  |  |  |  |  |  |  | <0.001 |
| 16–29 | 19.2 | 24.1 | 14.3 | 7.7 | 9.9 | 11.0 | 10.3 | 6.3 | 13.7 |  |
| 30–49 | 36.8 | 36.9 | 33.0 | 28.0 | 36.0 | 47.9 | 28.8 | 43.9 | 39.5 |  |
| 50–64 | 28.0 | 23.5 | 34.1 | 44.3 | 40.2 | 29.7 | 38.1 | 36.3 | 33.3 |  |
| 65+ | 16.0 | 15.5 | 18.7 | 20.0 | 13.8 | 11.4 | 22.8 | 13.5 | 13.5 |  |
| **Education, %** |  |  |  |  |  |  |  |  |  | <0.001 |
| No qualification | 11.4 | 11.1 | 13.1 | 13.2 | 12.5 | 7.9 | 12.8 | 14.1 | 10.5 |  |
| A-Level, GCSE, other | 52.3 | 52.1 | 53.5 | 54.5 | 52.5 | 50.7 | 52.9 | 52.2 | 52.1 |  |
| Degree or higher | 36.3 | 36.9 | 33.4 | 32.3 | 35.0 | 41.4 | 34.3 | 33.7 | 37.4 |  |
| **Ethnicity, %** |  |  |  |  |  |  |  |  |  | <0.001 |
| White | 93.6 | 92.7 | 94.5 | 96.5 | 94.6 | 94.7 | 96.0 | 95.2 | 95.2 |  |
| Black | 1.6 | 1.9 | 1.3 | 0.5 | 1.6 | 1.3 | 0.9 | 1.3 | 1.4 |  |
| Indian | 1.7 | 2.0 | 1.5 | 0.9 | 1.3 | 1.2 | 0.7 | 1.3 | 0.9 |  |
| Pakistani/Bangladeshi | 1.2 | 1.2 | 1.4 | 1.2 | 1.4 | 1.2 | 1.3 | 1.4 | 1.3 |  |
| Other Asian/other | 1.8 | 2.2 | 1.4 | 0.9 | 1.1 | 1.7 | 1.0 | 0.8 | 1.2 |  |
| **Occupational class, %** |  |  |  |  |  |  |  |  |  | <0.001 |
| Not employed | 39.0 | 38.2 | 41.2 | 45.1 | 40.4 | 29.2 | 44.8 | 45.2 | 36.2 |  |
| Management & professional | 28.0 | 29.1 | 24.5 | 22.5 | 27.6 | 34.1 | 23.8 | 24.6 | 28.3 |  |
| Intermediate | 14.0 | 14.0 | 14.0 | 14.2 | 13.2 | 15.5 | 12.7 | 13.2 | 15.4 |  |
| Routine | 18.9 | 18.7 | 20.3 | 18.1 | 18.8 | 21.3 | 18.7 | 17.0 | 20.1 |  |
| **Income quintile, %** |  |  |  |  |  |  |  |  |  | <0.001 |
| 1 (low) | 14.9 | 14.8 | 16.9 | 15.1 | 13.9 | 13.6 | 13.2 | 16.0 | 16.2 |  |
| 2 | 18.3 | 17.3 | 19.6 | 21.1 | 23.2 | 18.0 | 20.3 | 21.0 | 19.6 |  |
| 3 | 19.2 | 19.2 | 19.6 | 17.7 | 19.5 | 19.4 | 17.8 | 22.3 | 17.6 |  |
| 4 | 21.8 | 22.1 | 20.9 | 22.0 | 20.3 | 21.7 | 23.4 | 19.1 | 20.8 |  |
| 5 (high) | 25.8 | 26.7 | 23.0 | 24.1 | 23.1 | 27.3 | 25.3 | 21.6 | 25.8 |  |
| **Working status, %** |  |  |  |  |  |  |  |  |  | <0.001 |
| Not in paid employment | 35.6 | 34.3 | 38.1 | 42.5 | 38.7 | 27.2 | 42.1 | 42.9 | 33.1 |  |
| Full-time | 47.4 | 49.5 | 43.6 | 39.1 | 40.9 | 56.0 | 40.4 | 40.2 | 45.3 |  |
| Part-time | 17.0 | 16.3 | 18.3 | 18.4 | 20.4 | 16.8 | 17.4 | 16.9 | 21.6 |  |
| **Children in household, %** |  |  |  |  |  |  |  |  |  | <0.001 |
| None | 72.7 | 71.9 | 74.5 | 78.8 | 74.2 | 67.9 | 80.3 | 70.1 | 70.5 |  |
| 1 | 12.5 | 13.0 | 11.4 | 9.7 | 11.0 | 13.5 | 10.4 | 12.4 | 12.9 |  |
| 2 | 11.1 | 11.5 | 9.8 | 7.5 | 11.7 | 14.6 | 6.6 | 12.2 | 12.2 |  |
| 3+ | 3.7 | 3.6 | 4.2 | 4.0 | 3.1 | 3.9 | 2.7 | 5.3 | 4.4 |  |
| Married or cohabiting, % | 66.0 | 62.5 | 69.1 | 74.1 | 68.5 | 74.9 | 69.8 | 75.8 | 71.6 | <0.001 |
| Fair or poor self-rated health, % | 15.8 | 14.7 | 19.2 | 17.9 | 20.6 | 14.1 | 16.6 | 21.6 | 16.4 | <0.001 |
| **Household size, %** |  |  |  |  |  |  |  |  |  | <0.001 |
| 1 | 13.8 | 15.1 | 13.1 | 12.2 | 13.3 | 11.1 | 13.8 | 8.0 | 9.3 |  |
| 2 | 35.7 | 33.4 | 39.1 | 44.6 | 38.3 | 35.0 | 43.8 | 37.9 | 37.9 |  |
| 3–4 | 40.2 | 41.0 | 37.5 | 34.4 | 38.2 | 44.1 | 34.7 | 42.9 | 41.3 |  |
| 5+ | 10.3 | 10.5 | 10.3 | 8.8 | 10.2 | 9.9 | 7.7 | 11.1 | 11.5 |  |
| **Wave outcome observed, %** |  |  |  |  |  |  |  |  |  | <0.001 |
| 7 | 7.4 | 8.4 | 7.8 | 2.9 | 0.2 | 0.0 | 9.1 | 10.1 | 2.7 |  |
| 9 | 6.3 | 6.8 | 4.9 | 5.6 | 6.1 | 3.0 | 6.4 | 6.3 | 6.6 |  |
| 11 | 7.5 | 7.4 | 7.8 | 9.6 | 6.3 | 8.6 | 7.9 | 6.2 | 6.8 |  |
| 13 | 78.8 | 77.4 | 79.5 | 81.9 | 87.4 | 88.3 | 76.7 | 77.4 | 83.8 |  |
| Age at baseline, mean (SD) | 46.5 (17.1) | 44.5 (17.9) | 49.6 (16.2) | 53.1 (14.2) | 49.5 (14.6) | 47.0 (13.8) | 52.5 (15.7) | 49.8 (12.9) | 47.5 (14.7) | <0.001 |
| GHQ at baseline, mean (SD) | 11.0 (5.3) | 10.8 (5.2) | 11.2 (5.4) | 11.5 (5.5) | 11.5 (5.5) | 11.3 (5.3) | 11.2 (5.4) | 12.0 (5.8) | 11.2 (5.1) | <0.001 |
| SF-12 at baseline, mean (SD) | 50.6 (10.4) | 51.1 (10.2) | 48.9 (11.1) | 49.5 (10.9) | 49.6 (10.7) | 51.4 (9.7) | 49.8 (10.8) | 49.2 (10.9) | 49.7 (10.5) | <0.001 |

Recurrent caregivers were predominantly women (60.2%) and concentrated in the 30 to 49 and 50 to 64 age groups. They were less likely to be physically inactive than non-caregivers (48.5% vs 54.2%), reported slightly higher fruit and vegetable consumption (3.8 vs 3.6 portions per day), and had a lower prevalence of problematic drinking (40.9% vs 48.5%) but a higher prevalence of smoking (17.4% vs 11.8%).

### Associations between caregiving trajectories and health behaviours

#### Physical inactivity

After adjustment, recurrent caregivers had lower odds of physical inactivity than non-caregivers (OR=0.65, 95%CI: 0.53/0.81), and no other caregiving classes were associated with physical inactivity in the adjusted models. In unadjusted models, Former-long and Long-term caregivers had higher odds of physical inactivity, but these associations attenuated after adjustment for confounders. The association was modified by sex (Wald p=0.05). In sex-stratified models, Former-long caregiving was associated with higher odds of physical inactivity among men (OR 1.44, 95% CI: 1.10/1.87) but not women (OR 0.96, 95% CI 0.79/ 1.16), whereas Long-term caregiving was associated with lower odds among women (OR 0.83, 95% CI 0.70 to 0.98) but not men (OR 1.14, 95% CI: 0.89/1.44). Recurrent caregivers had lower odds in both sexes.

#### Fruit and vegetable consumption

Several trajectories were associated with higher fruit and vegetable consumption in unadjusted models, but these associations attenuated after adjustment for baseline intake and covariates, and no trajectory was associated with consumption in the fully adjusted model. Associations differed by age group (p for interaction = 0.01). In the youngest group (16 to 29 years), Emerging-long caregivers consumed fewer daily portions than non-caregivers (coefficient −0.55, 95% CI: −0.92/−0.18), whereas in the oldest group (65 years and over), Recurrent (coefficient 0.52, 95% CI: 0.06/0.98) and Former-long caregivers (coefficient 0.31, 95% CI 0.04/0.58) consumed more fruit and vegetables (Figure 4).

#### Problematic drinking

In unadjusted models, all caregiving classes had lower odds of problematic drinking than non-caregivers, but these associations attenuated after adjustment for confounders. Recurrent caregivers retained lower odds after adjustment (OR=0.75, 95%CI: 0.59/0.94), and Long-term caregivers showed lower odds in the same direction, although the interval included the null (OR=0.87, 95%CI: 0.74/1.01). No other class was associated after adjustment, and no interactions by sex or age were detected.

#### Smoking

Recurrent caregivers were the only class associated with smoking after full adjustment, with higher odds than non-caregivers (OR=1.67, 95%CI: 1.17/2.40). The association persisted after adjustment for baseline smoking status, indicating higher odds of smoking at follow-up among Recurrent caregivers than among non-caregivers with the same baseline status. There was no evidence of effect modification by sex or age group.

**Figure 2:**
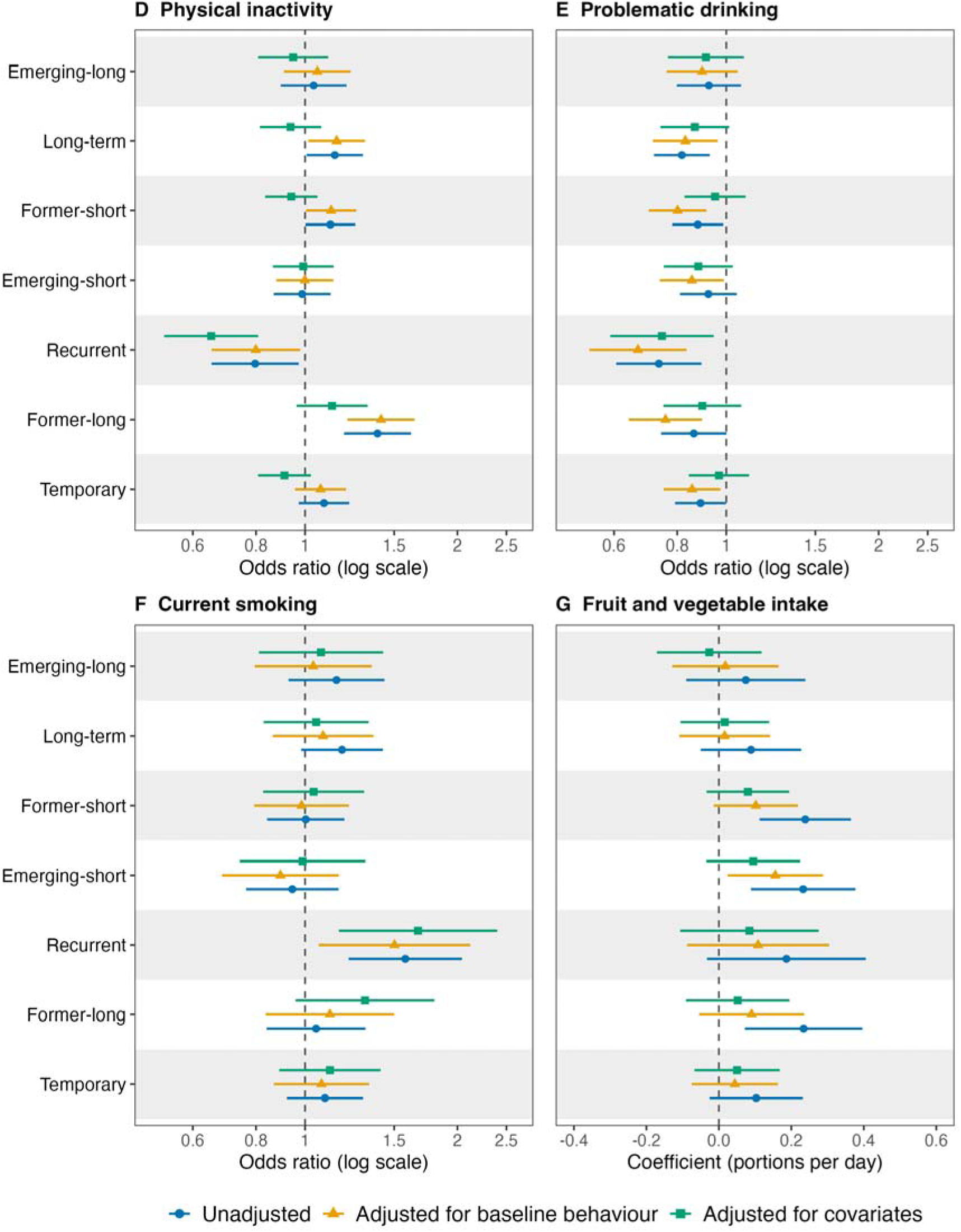
Associations between unpaid caregiving trajectory class and four health behaviours. The four panels show physical inactivity (D), problematic drinking (E), current smoking (F) and fruit and vegetable intake (G). Points are the association for each latent caregiving trajectory class compared to non-caregivers as reference category and horizontal lines are 95% confidence intervals. For physical inactivity, problematic drinking and current smoking the estimates are odds ratios plotted on a log scale, with the dashed line at 1. For fruit and vegetable intake the estimates are linear regression coefficients [in portions per day], with the dashed line at 0. Three models are shown for each class. The unadjusted model is marked with circles, the model additionally adjusted for the baseline level of the same behaviour with triangles, and the model adjusted for selected covairates with squares. The fully adjusted model accounts for baseline behaviour, age, sex, education, ethnicity, occupational class, income, employment, presence of children, cohabitation, household composition, psychological distress, self-rated health [and the SF-12 physical component for physical inactivity], and the survey wave of outcome measurement. All estimates are survey weighted and pooled across [m = 10] multiply imputed datasets using Rubin’s rules. Axis ranges differ between panels. OR=odds ratio; CI=confidence interval.

**Figure 3:**
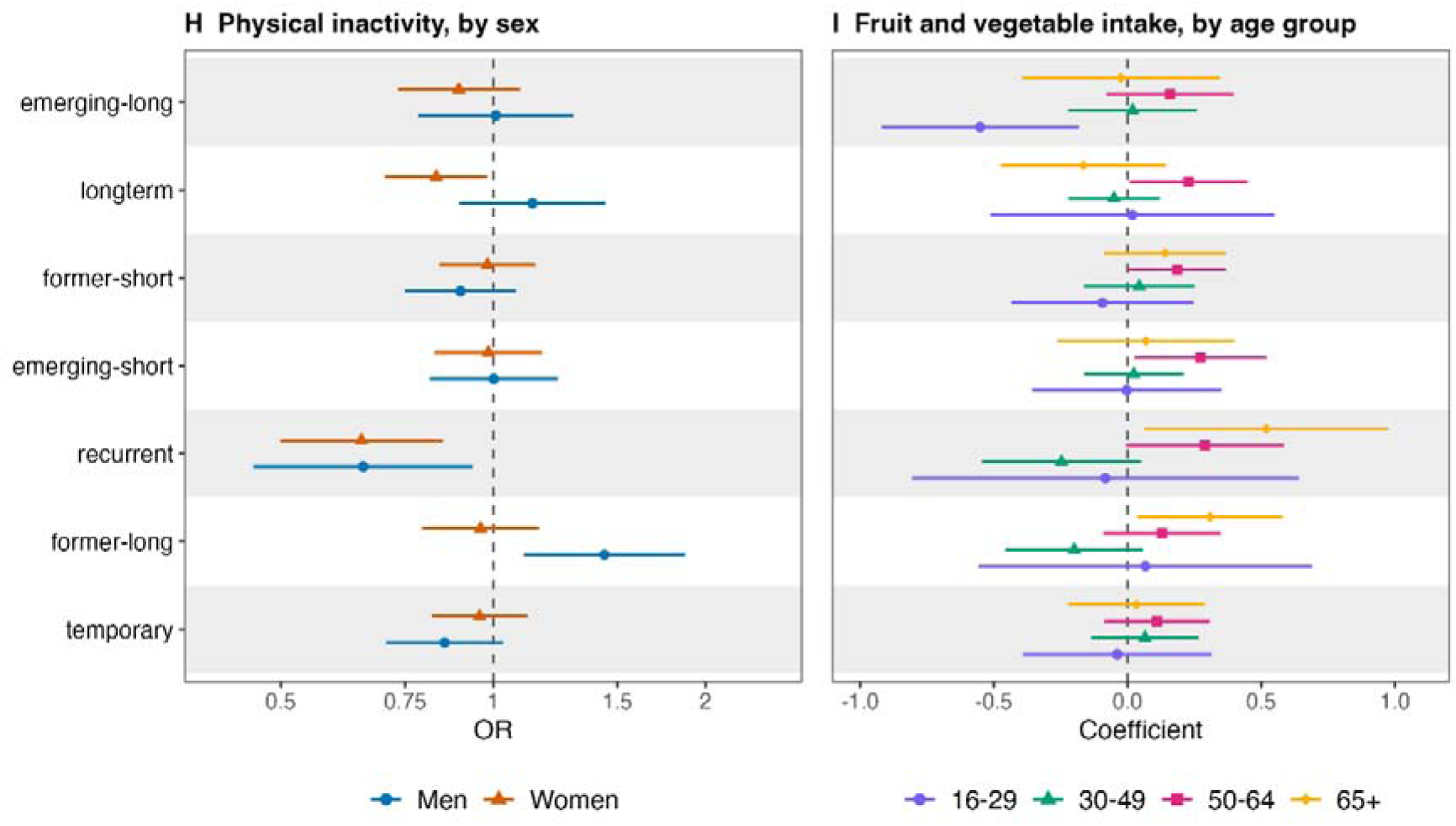
Stratified associations between caregiving trajectory class and health behaviours in subgroups with evidence of effect modification. Fully adjusted estimates are shown within strata for the two outcome and modifier combinations where we found evidence of effect modification. Panel H shows physical inactivity by sex, with odds ratios for men (circles) and women (triangles) plotted on a log scale and the dashed line at 1. Panel I shows fruit and vegetable intake by age group, with linear regression coefficients [in portions per day] for ages 16-29 (circles), 30-49 (triangles), 50-64 (squares) and 65+ (diamonds) and the dashed line at 0. Points are estimates for each latent caregiving trajectory class relative to non-caregivers as reference class within each stratum, and horizontal lines are 95% confidence intervals. Each model adjusts for the health behaviour at baseline and selected covairates. Estimates are survey weighted and pooled across [m = 10] multiply imputed datasets. Axis ranges differ between panels. OR=odds ratio; CI=confidence interval.

### Sensitivity analyses

Sequence analysis and an observed-transitions comparator, were broadly consistent with the main findings and are reported in Supplement 10 and 11.

## Discussion

### Summary of findings

Using latent class analysis of 12 waves of nationally representative panel data, we identified eight caregiving trajectories differing in the timing, duration, and recurrence of care (H1). The Recurrent class, although the smallest among caregivers, showed the most consistent associations, and these only partly matched our hypotheses. Recurrent caregivers had higher odds of smoking, in line with H2, but lower odds of physical inactivity, contrary to H2, and lower odds of problematic drinking, consistent with H3. Contrary to H4, associations were not less favourable among women, and longer-duration caregiving was more strongly associated with inactivity in men. H5 was partially supported, with age differences confined to fruit and vegetable consumption.

### Interpretation of findings

That recurrent caregivers were more physically active than non-caregivers may appear counterintuitive. However, unpaid care frequently involves physical tasks such as personal care, mobility assistance, and household management, which may raise incidental activity.^6,27^ The pattern is consistent with the physical activity paradox, whereby occupational or task-related activity does not confer the cardiovascular and mortality benefits of leisure-time activity and in some studies is linked to adverse outcomes.^28,29^ Lower physical inactivity may therefore reflect the physical burden of care rather than a health-promoting choice, and further evidence is needed.

Recurrent caregivers were the only class associated with smoking after adjustment. This adds to longitudinal evidence which found that transitioning into higher-intensity caregiving increased the probability of smoking.^11^ The elevated odds of smoking among recurrent caregivers are consistent with the stress process model, in which caregiving burden promotes maladaptive coping.^5^

Caution is also warranted in interpreting lower problematic drinking as a health behaviour choice, as care demands may limit social opportunities and require caregivers to remain vigilant to the care recipient’s needs^6,30^ A second possibility is a healthy carer selection effect, whereby individuals able to take on and relinquish care repeatedly have lower baseline consumption.^31^ Our finding is consistent with cross-sectional studies reporting lower alcohol intake among caregive,^30,32^ although the two most robust longitudinal population studies found no difference in alcohol consumption between caregivers and non-caregivers.^9,10^

Associations with fruit and vegetable consumption were concentrated in the youngest age group, where Emerging-long caregivers reported around half a portion per day less than non-caregivers. Taking on longer-duration care in early adulthood may coincide with other transitions such as leaving education or entering employment, at a stage when dietary habits are still forming. Associations were weaker elsewhere, including the higher intake among Recurrent and Former-long caregivers in late adulthood. This pattern is consistent with a comparable longitudinal study, where transitioning into care was not associated with a change in non-daily fruit and vegetable consumption except among co-resident men.^9^ If caregiving affects diet through stress pathways, fruit and vegetable intake may not be an optimal measure, and fast food consumption or snacking may be more informative.^33^

Evidence that associations varied by age or sex was limited, which is somewhat surprising given the strongly gendered nature of unpaid care.^5,16^ This may indicate that behavioural associations are broadly consistent across these groups, but it may also reflect limited power to detect interactions, since several classes were small. However, the composition of the Recurrent class should be acknowledged, as these caregivers were predominantly women and concentrated in mid-life, meaning the absolute burden of the associations reported here falls disproportionately on this group.

This study also makes a methodological contribution. Few studies have used latent class or related trajectory methods to capture how caregiving accumulates across time.^34^ A recent analysis of the Health and Retirement Study applied such an approach and likewise identified a recurrent caregiving class, though it drew on retrospective life-history reports and named recall bias in the timing and duration of care episodes as a key limitation.^35^ Our use of 12 waves of prospectively collected annual data avoids this source of bias, though at the cost of a shorter observation window that captures caregiving only within the study period rather than across the full lifecourse.

### Implications

Social care policy in the UK and elsewhere is largely designed around the assumption that caregiving is a single occassion with support often tied to hours of care currently provided.^36,37^ The identification of distinct trajectory classes, each with different implications for health, suggests that policy should also account for the episodic nature of care. Recurrent caregivers in particular should be considered a target group for smoking cessation. Because the elevated smoking likely reflects stress-related coping, downstream behaviour change interventions alone may be insufficient, and upstream measures addressing the structural causes of caregiving stress, including respite, financial support, and flexible employment, are also warranted.^4,38^

### Strengths and limitations

Several limitations warrant consideration. All measures, including caregiving status and the four health behaviours, were self-reported and are therefore subject to recall and social desirability bias. By wave 13 the study had lost 64.5% of its initial sample, although attrition patterns are comparable to those observed in other longitudinal studies.^39^ The physical inactivity measure does not distinguish occupational from leisure activity, and it remains unclear whether lower inactivity in some classes translates into better health outcomes. Physical inactivity and problematic drinking were dichotomised against validated thresholds, which entails a loss of information and may obscure gradual changes that do not cross those thresholds. Further, because the baseline health behaviour questions differed from those used at follow-up, adjustment for prior behaviour may be prone to residual confounding. However, we preferred this to omitting baseline adjustment or confining the trajectories to waves 7 to 13. The entropy of the selected model (0.74) indicates moderate class separation and therefore some misclassification, although values above 0.60 are generally considered acceptable when theoretically grounded,^40^ and several classes were small. However, strengths include a large, nationally representative panel of 25,049 participants with up to 12 years of annual follow-up, trajectories derived empirically rather than imposed through researcher- defined categories, and sensitivity analyses using alternative classification methods. As with all observational studies, causality cannot be assumed.

## Conclusion

Using latent class analysis of 12 waves of nationally representative panel data, we identified eight caregiving trajectories, comprising temporary, former-short, former-long, emerging-short, emerging-long, long-term, and recurrent caregivers alongside a sustained non- caregiving class. Recurrent caregivers showed the most consistent associations with health behaviours, with higher odds of current smoking and lower odds of physical inactivity and problematic drinking relative to non-caregivers, and no association with fruit and vegetable consumption. Caution is warranted in interpreting these health-promoting behaviours as healthy lifestyle choices, since they may instead mask the burden of care. Recurrent caregiving should be recognised by researchers and practitioners as a further dimension of caregiving alongside its intensity and duration, and support systems should be better equipped for people who move repeatedly into and out of the caregiving role.

## Declaration of Competing Interests

The authors declare no competing interests. September 2026

## Ethical approval

The project uses secondary data publicly available from the UK Data Service (https://ukdataservice.ac.uk) and Ethical approval for the Understanding Society main study and Innovation Panel was granted by the University of Essex Ethics Committee. Therefore, ethical approval for this project was not required.

## Data Availability Statement

This study uses data from the UK Household Longitudinal Study (UKHLS), which is publicly available through the UK Data Service (https://ukdataservice.ac.uk) under standard access conditions. The analytical code used for this study can be made available through github upon acceptance of the paper for publication.

## Funding

This project is funded by the UK Economic and Social Research Council (ESRC) through the UBEL-DTP (UCL, Bloomsbury and East London Doctoral Training Partnership) and is registered under the grant reference ES/P000592/1. The funder was not involved in project design, analysis or write up of findings. AM and BX were supported by funding though the Joint Programming Initiative More Years, Better Lives (JPI MYBL) from the UK Economic and Social Research Council (ES/W001454/1). AM, BX and EP are supported by UK Economic and Social Research Council funding for *Equalise*: ESRC Centre for Lifecourse Health Equity (ES/Z504270/1).

## Notes

### Competing Interest Statement

The authors have declared no competing interest.

