## Supplement for "Caregiving transition trajectories and health behaviours: a latent class analysis of the UK Household Longitudinal Study"

### S1: Variable definition

**Table S.1:** Variable description; UKHLS, UK Household Longitudinal Study; AUDIT-C, Alcohol Use Disorders Identification Test - Consumption; NS-SEC, National Statistics Socio-economic Classification; OECD, Organisation for Economic Co-operation and Development; GHQ-12, 12-item General Health Questionnaire; SF-12 PCS, 12-item Short Form Survey Physical Component Summary

| Variable | Definition |
|---|---|
| ***Outcomes*** | |
| Physical inactivity | Binary variable derived from questions on vigorous and moderate activity in the past 7 days, which align with the International Physical Activity Questionnaire. Participants were classified as physically active (=0) if they reported ≥75 minutes of vigorous activity, ≥150 minutes of moderate activity, or ≥150 minutes of moderate and vigorous activity combined per week, in line with UK Chief Medical Officer recommendations; otherwise as physically inactive (=1). Walking was excluded because the UKHLS item captures all walking of at least 10 minutes, including walking to work, whereas the Chief Medical Officer definition of moderate activity refers to brisk walking only. |
| Fruit and vegetable consumption | Continuous variable measuring average daily portions of fruit and vegetables, derived from frequency and portion size items. Not dichotomised because of the near-normal distribution and the absence of consensus on a cut-off. |
| Smoking | Binary variable based on the smoking status item. Current smoker (=1) or non-smoker (=0). |
| Problematic drinking | Binary variable derived from the three-item AUDIT-C. Problematic drinking (=1) defined as a score ≥3 (women) or ≥4 (men); otherwise no problematic drinking (=0). |
| ***Health behaviours at baseline*** | |
| Walking at baseline | Categorical proxy for physical activity at baseline, as the physical activity module in waves 2 and 5 covered walking only. Derived from the number of days in the past four weeks on which participants walked for 30 minutes or more: none; 1–2 days; 3–4 days; 5–6 days; every day. A sports-based alternative measure was not used because of 38% missingness. |
| Fruit and vegetable consumption at baseline | Categorical variable derived from three items in waves 2 and 5 on days per week eating fruit, days per week eating vegetables, and portions consumed on a typical day: zero portions; 1–3 portions; 4 portions; 5 or more portions. |
| Alcohol consumption at baseline | Categorical proxy for problematic drinking at baseline, as AUDIT-C was not administered in waves 2 and 5. Derived from items on whether participants had ever had an alcoholic drink and how often they drank in the past 12 months: no drinks; monthly but less than weekly; 1–4 drinks per week; 5 or more drinks per week. |
| Smoking at baseline | Categorical variable based on smoking status reported at the baseline wave. |

| Variable | Definition |
|---|---|
| ***Covariates*** | |
| Sex | Binary variable: male or female. |
| Age / age group | Continuous variable, also grouped into 16–29, 30–49, 50–64 and 65+ years to reflect life course stages. |
| Ethnicity | Categorical variable: white; black; Indian; Pakistani or Bangladeshi; other Asian or other ethnicity. |
| Cohabiting status | Binary variable: cohabiting (married or partnered) or non-cohabiting (single, divorced, separated, widowed). |
| Household size | Categorical variable: 1-person; 2-person; 3–4 person; 5+ person household. |
| Number of children | Categorical variable: 0; 1; 2; 3 or more children under 16 living in the household. |
| Education | Categorical variable based on highest qualification: no qualification; GCSE, A-level or other qualification; degree or higher qualification. |
| Employment status | Categorical variable: full-time (≥30 hours); part-time (<30 hours); not in paid employment. |
| Occupational class | Categorical variable (NS-SEC): managerial or professional; intermediate; routine or manual; not employed. |
| Household income quintiles | Categorical variable derived from net household income adjusted using the OECD equivalence scale. |
| Self-rated general health | Binary variable: fair or poor (=1); good, very good or excellent (=0). |
| GHQ-12 | Continuous variable (0–36) from the 12-item General Health Questionnaire measuring psychological distress. Higher scores indicate greater distress. |
| SF-12 PCS | Continuous variable (0–100) measuring physical health functioning. Higher scores indicate better functioning. |
| Baseline wave | Wave at which the participant was first observed. |
| Outcome wave | Wave at which the outcome was measured: wave 7, 9, 11 or 13 for physical inactivity, fruit and vegetable consumption and problematic drinking; waves 5 to 13 for smoking. Included in adjusted models to account for period effects. |
| Number of waves observed | Number of waves in which the participant was observed. Range 2–4 for physical inactivity, fruit and vegetable consumption and problematic drinking; 2–9 for smoking. |

### S2: DAG

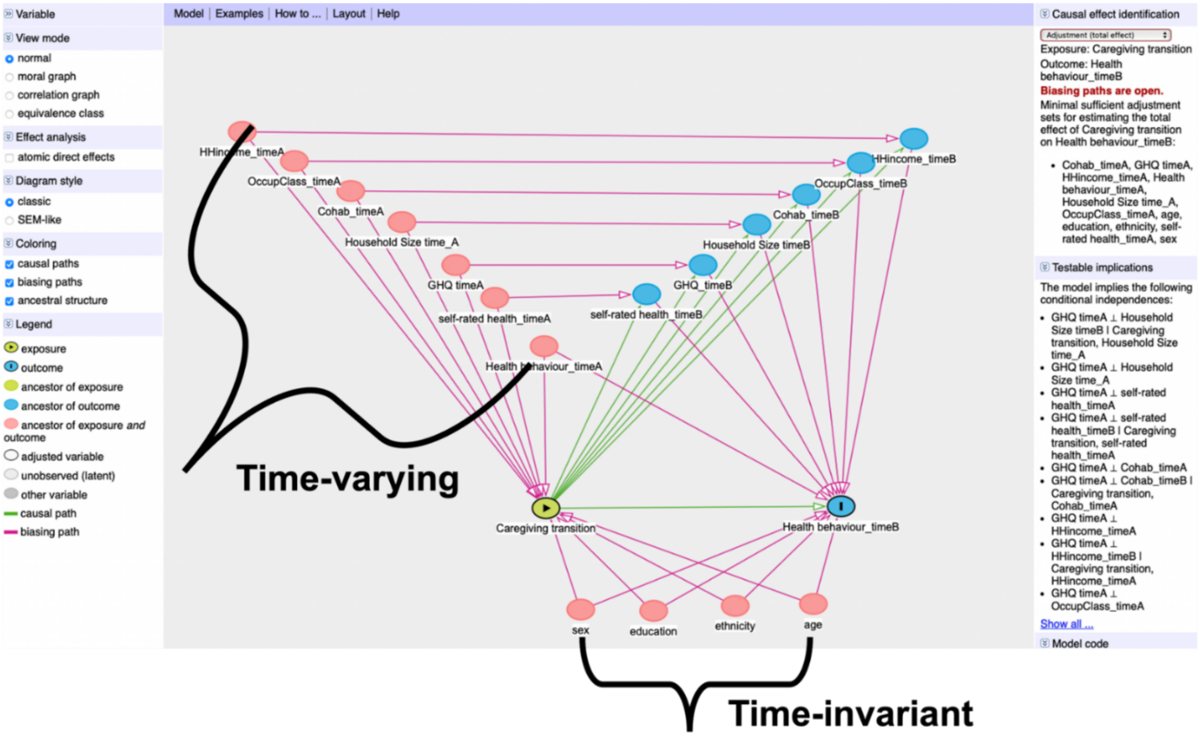

**Figure S.1:** Directed Acyclic Graph (DAG) illustrating assumed causal relationships between caregiving transitions and subsequent health behaviours. This DAG represents the hypothesised relationships between transitioning into or out of unpaid caregiving (exposure) and subsequent health behaviours (outcome). Time-varying covariates (measured at baseline [time A] and follow-up [time B]) include self-rated health, GHQ-12 score, income, occupational class, cohabitation status, and household size. Time-invariant covariates include sex, age, ethnicity, and education. Health behaviour at time A is included to account for baseline behaviour. Minimal sufficient adjustment for estimating the total causal effect of caregiving transition on health behaviour at time B includes: baseline values of cohabitation, GHQ-12, income, occupational class, household size, self-rated health, and health behaviour, along with time-invariant variables (age, sex, ethnicity, and education). Green arrows represent causal paths, pink lines represent open biasing paths, and black arrows indicate variables classified as time-varying. This model was created in DAGitty (Textor et al., 2016).

### S3: Conceptual framework

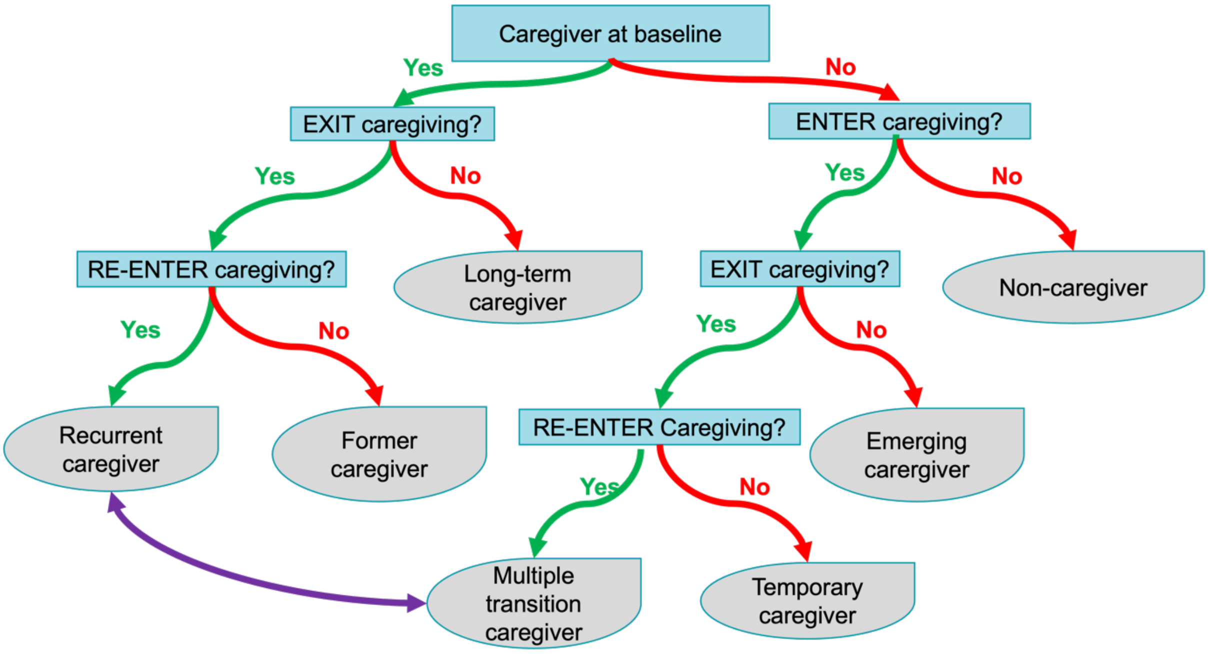

**Figure S.2:** Conceptual framework for classifying caregiving trajectories. Trajectories are defined by caregiving status at baseline and by subsequent exits from and re-entries into caregiving. Blue boxes denote decision points and grey ellipses the resulting trajectory types. Participants with more than one transition may be classified as recurrent or multiple-transition caregivers depending on the timing of re-entry.

#### S4: Sample Size

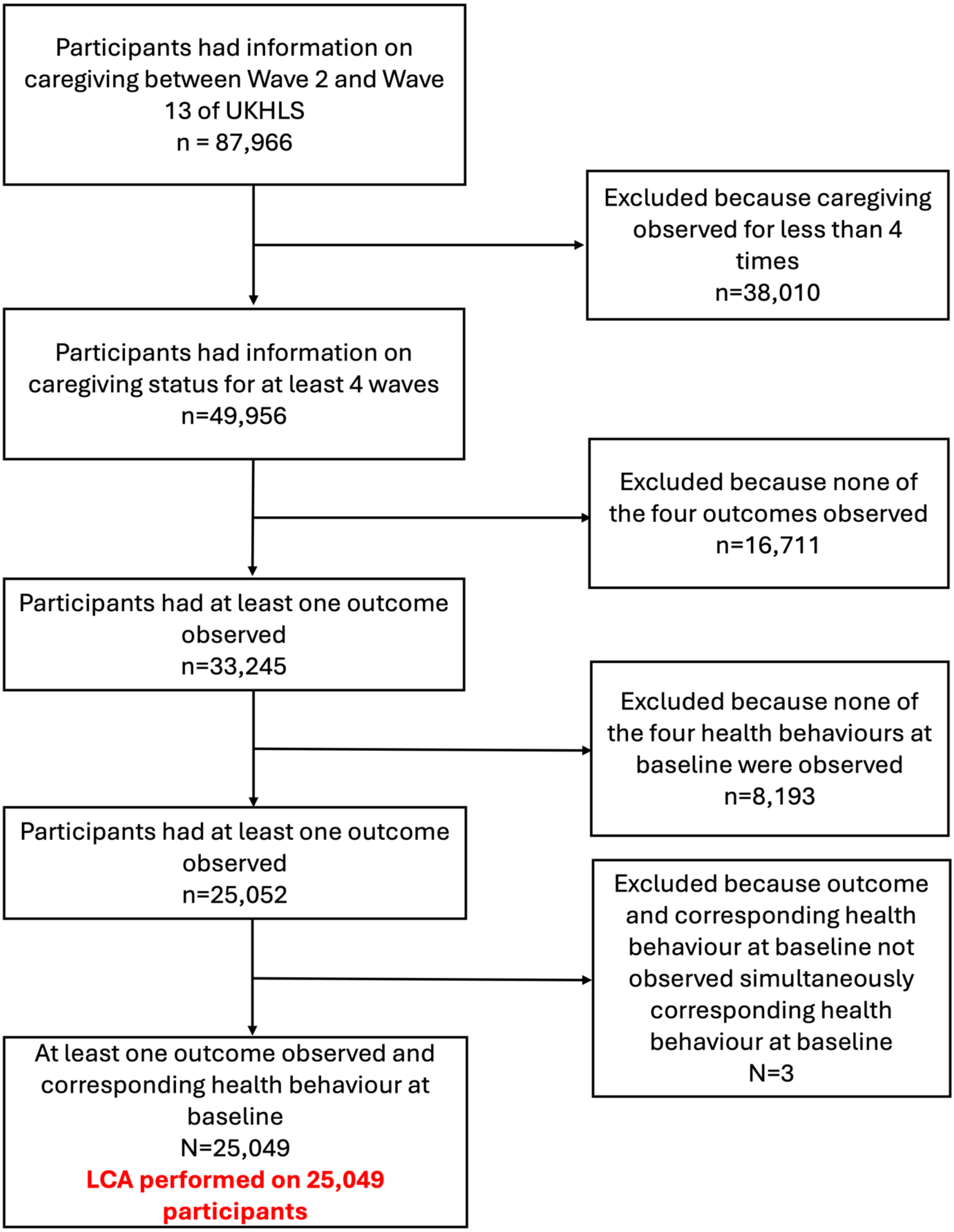

**Figure S.3:** Derivation of the analytic sample, UK Household Longitudinal Study waves 2 to 13. Latent class analysis was performed on all 25,049 eligible participants.

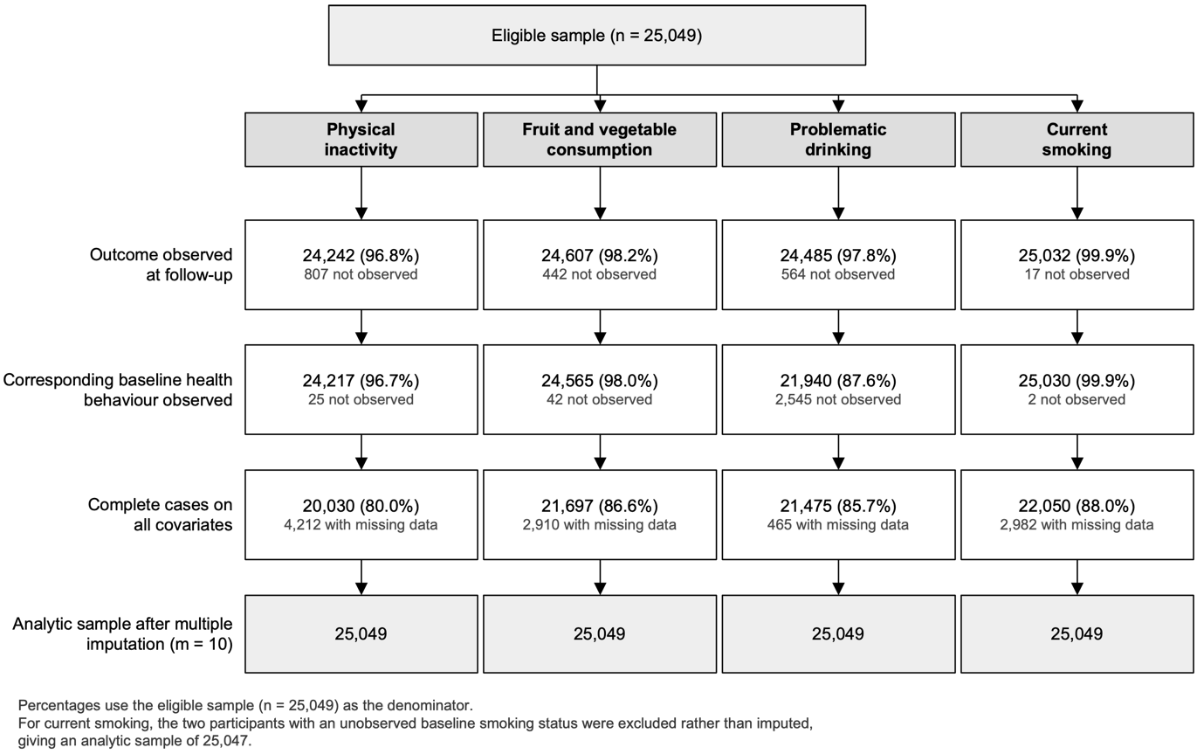

**Figure S. 4:** Figure S4. Observed data at each analytic stage by outcome, eligible sample (n = 25,049). Percentages use the eligible sample as the denominator. Counts at each stage are cumulative rather than sequential subtractions. All missing values were multiply imputed (m = 10) and analyses were conducted on the full eligible sample, except for current smoking, where two participants with unobserved baseline smoking status were excluded rather than imputed, giving an analytic sample of 25,047.

**Table S.2:** Missing data before multiple imputation, eligible sample (n = 25,049).
All counts and percentages use the eligible sample as the denominator; counts are not mutually exclusive across variables. All missing values were multiply imputed (m = 10) and substantive analyses were conducted on the full eligible sample. GHQ-12, self-rated health, SF-12 and baseline drinking frequency are drawn from the self-completion questionnaire and their missingness is largely concurrent: 2,161 participants (8.6%) were missing on exactly these four variables. GHQ-12, 12-item General Health Questionnaire; SF-12, 12-item Short Form Health Survey.

| Variable | n missing | % |
|---|---|---|
| **Outcome at follow-up** | | |
| Physical inactivity | 807 | 3.22 |
| Fruit and vegetable consumption | 442 | 1.76 |
| Problematic drinking | 564 | 2.25 |
| Current smoking | 17 | 0.07 |
| **Health behaviour at baseline** | | |
| Walking | 26 | 0.10 |
| Fruit and vegetable consumption | 45 | 0.18 |
| Drinking frequency | 2,675 | 10.68 |
| Smoking status | 2 | 0.01 |
| **Covariates** | | |
| Age | 0 | 0.00 |
| Sex | 0 | 0.00 |
| Education | 53 | 0.21 |
| Ethnicity | 10 | 0.04 |
| Occupational class | 195 | 0.78 |
| Income quintile | 28 | 0.11 |
| Employment status | 2 | 0.01 |
| Children in household | 0 | 0.00 |
| Cohabiting status | 4 | 0.02 |
| Household composition | 0 | 0.00 |
| GHQ-12 | 2,713 | 10.83 |
| Self-rated health | 2,495 | 9.96 |
| SF-12 physical functioning | 4,026 | 16.07 |

| Variable | n missing | % |
|---|---|---|
| **Missing data across all variables** | | |
| Complete on all variables | 19,274 | 76.95 |
| Missing on one or more variables | 5,775 | 23.05 |

### S5: Fit statistic

**Table S.3:** Matrix of average posterior probabilities for latent class assignment in the eight class solution across UKHLS waves 2 to 13 (n=25,049). Values represent the average probability of participants classified into each latent class (rows) being assigned to each possible class (columns). High diagonal values and low off-diagonal values indicate good classification quality.

| | [1] | [2] | [3] | [4] | [5] | [6] | [7] | [8] |
|---|---|---|---|---|---|---|---|---|
| [1] | **0.73** | 0.03 | 0.02 | 0.06 | 0.04 | 0.00 | 0.06 | 0.06 |
| [2] | 0.06 | **0.80** | 0.05 | 0.00 | 0.05 | 0.04 | 0.01 | 0.00 |
| [3] | 0.04 | 0.06 | **0.71** | 0.04 | 0.04 | 0.05 | 0.05 | 0.00 |
| [4] | 0.06 | 0.00 | 0.02 | **0.81** | 0.00 | 0.00 | 0.04 | 0.07 |
| [5] | 0.05 | 0.03 | 0.04 | 0.01 | **0.82** | 0.00 | 0.00 | 0.05 |
| [6] | 0.00 | 0.08 | 0.03 | 0.00 | 0.00 | **0.83** | 0.05 | 0.00 |
| [7] | 0.07 | 0.01 | 0.05 | 0.05 | 0.00 | 0.04 | **0.78** | 0.00 |
| [8] | 0.03 | 0.00 | 0.00 | 0.03 | 0.02 | 0.00 | 0.00 | **0.93** |

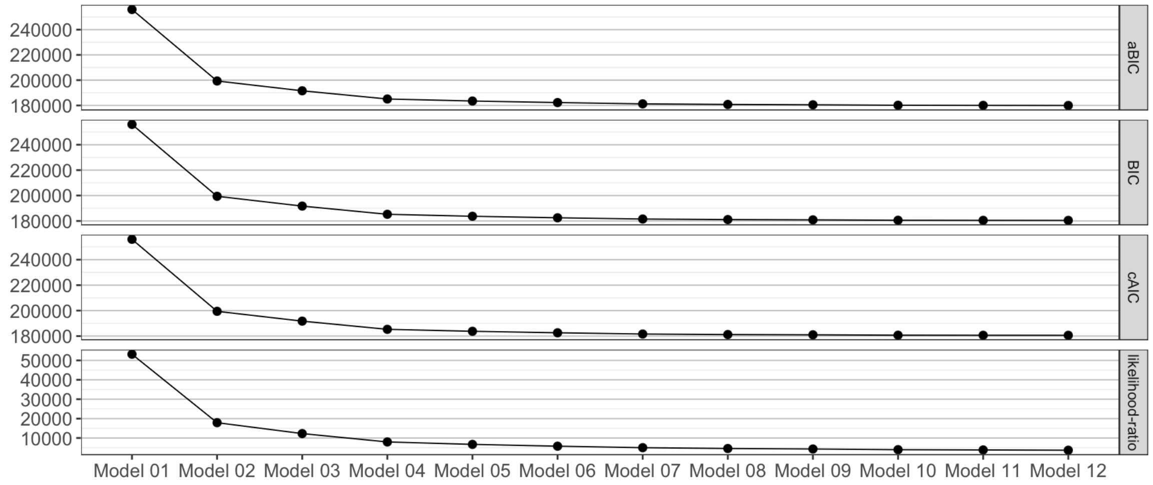

**Figure S.5:** Elbow Plot of model fit statistics for latent class analysis of caregiving status trajectories across UKHLS waves 2 to 13 (n=25,049). The plot displays fit indices (e.g., BIC, aBIC, cAIC) across different class solutions, with the 'elbow' indicating the optimal number of latent classes.

### S6: Posterior probabilities

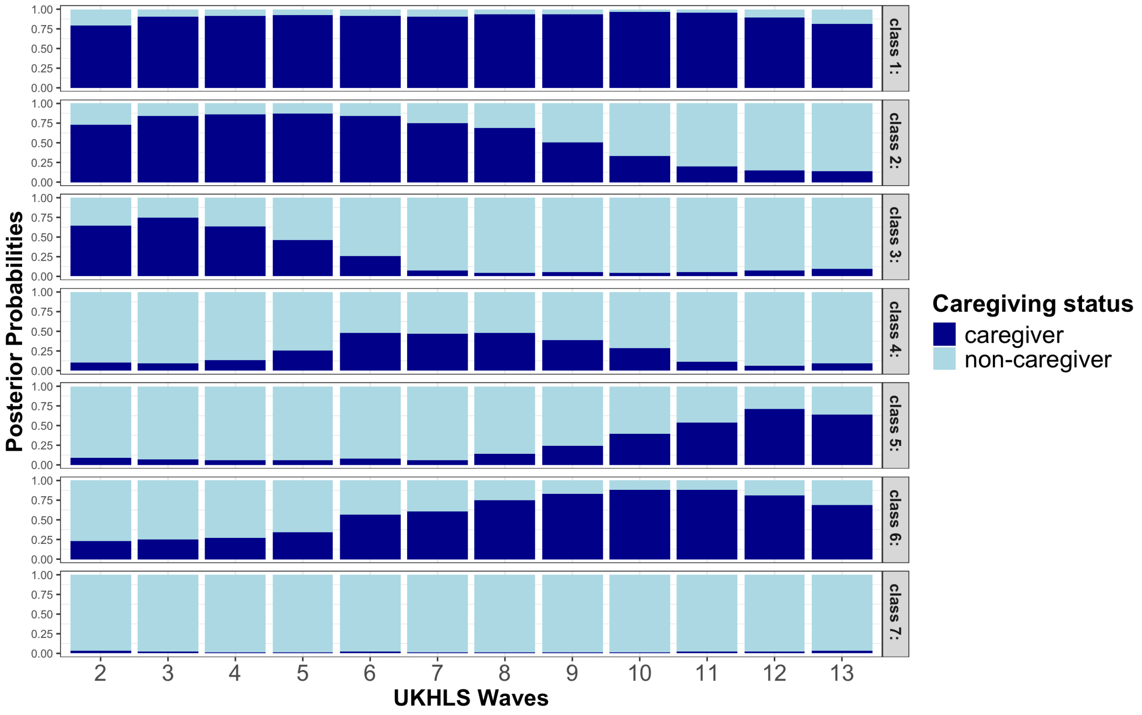

**Figure S.6:** Posterior probability for seven-class solution across UKHLS waves 2 to 13 (n=25,049). Each panel represents a latent class, showing the distribution of caregiving status over time.

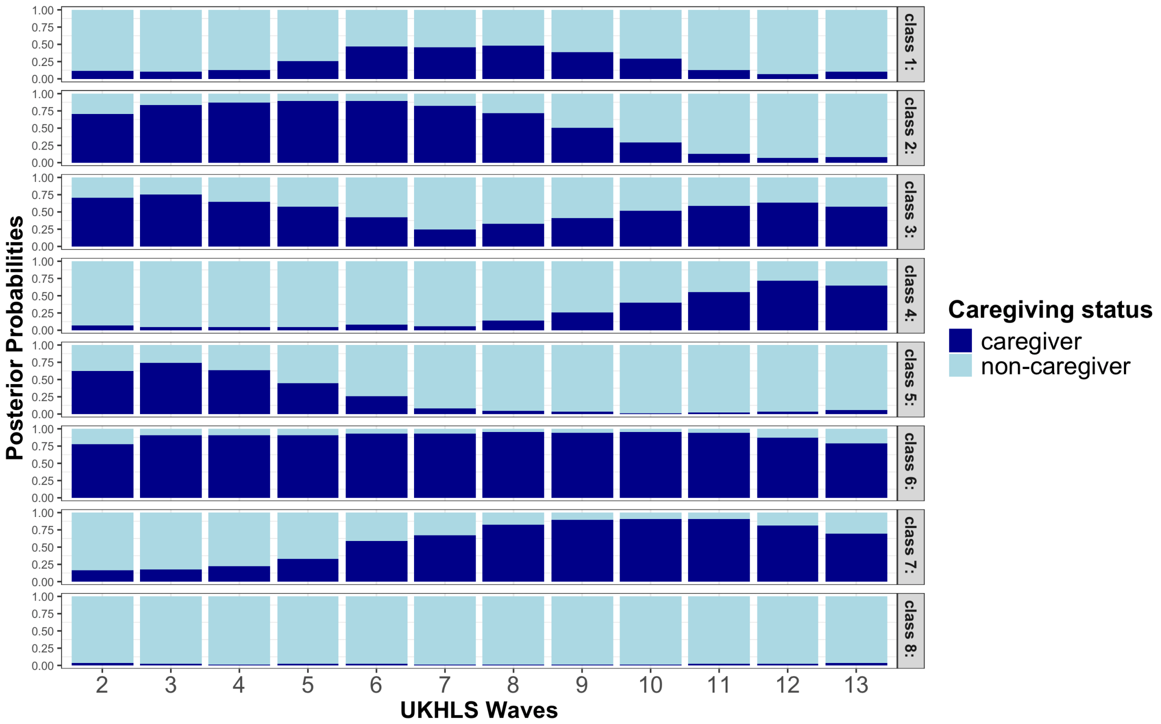

**Figure S.7:** Posterior probability for eight-class solution across UKHLS waves 2 to 13 (n=25,049). Each panel represents a latent class, showing the distribution of caregiving status over time.

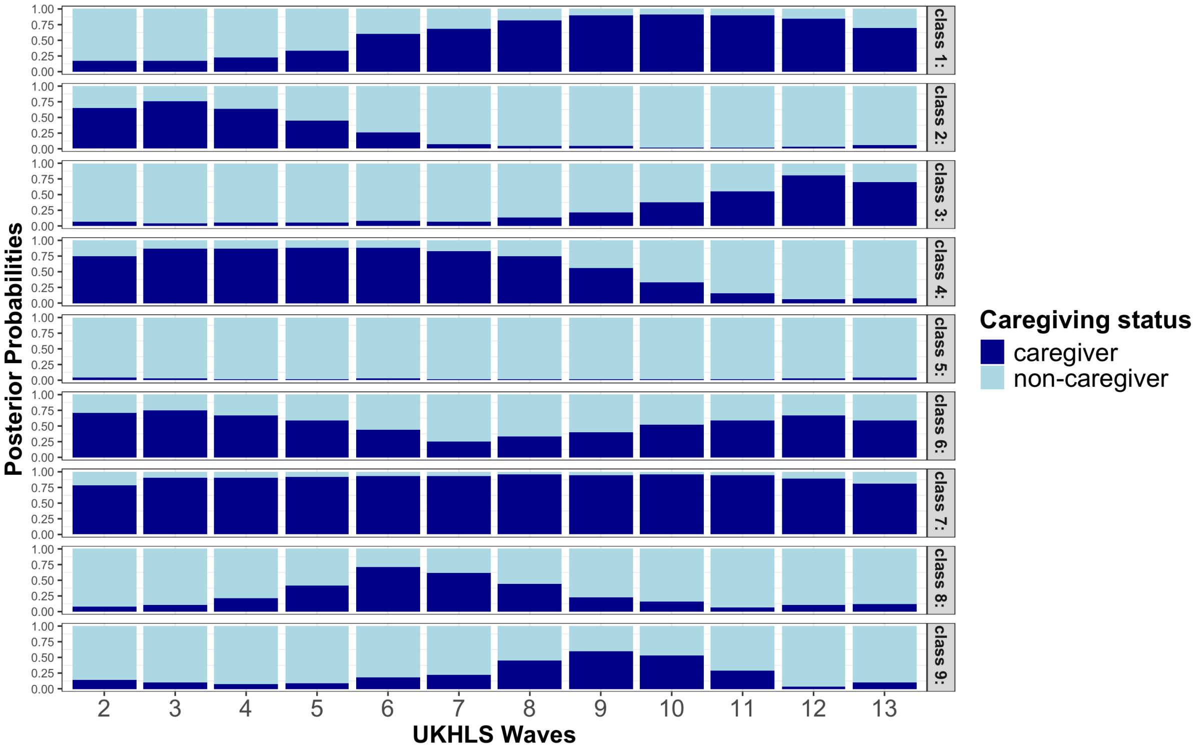

**Figure S.8:** Posterior probability for nine-class solution across UKHLS waves 2 to 13 (n=25,049). Each panel represents a latent class, showing the distribution of caregiving status over time.

### S7: Class definitions

**Table S.4:** Labels and definitions for latent classes identified through latent class analysis of caregiving status trajectories across UKHLS waves 2 to 13 (n=25,049). Latent classes are based on patterns of caregiving transitions over time.

| Class | Label | Definition |
|---|---|---|
| Class 1 | Temporary caregiver | Non-caregiver at start of the study, transition into caregiving and exit before last observation. |
| Class 2 | Former-long caregiver | Caregiver at start of study with a longer caregiving period prior exit (and short duration being non-caregiver) prior to last observation. |
| Class 3 | Recurrent caregiver | Caregiving at baseline with longer period of non-caregiving followed by a transition back to caregiving. |
| Class 4 | Emerging-short caregiver | Non-caregiver at start of study with longer period of non-caregiving followed by transition into care and short caregiving period until end of observation. |
| Class 5 | Former-short caregiver | Caregiver at start of study with a shorter caregiving period prior exit (and longer duration being non-caregiver) prior to last observation. |
| Class 6 | Long-term caregiver | Predominantly care-giver throughout observation period with occasional periods of non-caregiving. |
| Class 7 | Emerging-long caregiver | Non-caregiver at start of study with shorter period of non-caregiving followed by transition into care and longer caregiving period until end of observation. |
| Class 8 | Non-caregiver | Predominantly non-caregivers throughout observation period with some, rare short-term transition into care.. |

### S8: Full result table

**Table S.5:** Regression results for LCA for physical inactivity; logistic regression models predicting physical inactivity across latent caregiving intensity classes among UKHLS participants (n=25,049), showing pooled Odds Ratios from multiple imputation (m=10) and accounting for complex survey design and household-level clustering. Results are shown for three models: PA1a (unadjusted), PA2a (adjusted for walking at baseline), and PA3a (adjusted for selected covariates).

| | | **Model PA1a** | | **Model PA2a** | | **Model PA3a** | |
|---|---|---|---|---|---|---|---|
| | **Physical inactivity** | **Odds Ratio** | **95% CI** | **Odds Ratio** | **95% CI** | **Odds Ratio** | **95% CI** |
| Latent Class | No care | 1.00 | - | 1.00 | - | 1.00 | - |
| | Temporary | 1.08 | (0.96, 1.21) | 1.06 | (0.95, 1.19) | 0.91 | (0.81, 1.03) |
| | Former long | 1.41 | (1.21, 1.64) | 1.43 | (1.23, 1.66) | 1.13 | (0.96, 1.33) |
| | Recurrent | 0.8 | (0.65, 0.97) | 0.8 | (0.65, 0.97) | 0.65 | (0.53, 0.81) |
| | Emerging-short | 0.99 | (0.87, 1.13) | 1 | (0.88, 1.14) | 0.99 | (0.86, 1.14) |
| | Former-short | 1.12 | (1, 1.25) | 1.12 | (1, 1.26) | 0.94 | (0.83, 1.06) |
| | Long-term | 1.16 | (1.02, 1.33) | 1.17 | (1.03, 1.34) | 0.94 | (0.81, 1.08) |
| | Emerging long | 1.04 | (0.9, 1.2) | 1.06 | (0.91, 1.23) | 0.95 | (0.81, 1.11) |
| Walking at baseline | 0 days | | | 1.00 | - | 1.00 | - |
| | 1-2 days | | | 0.59 | (0.55, 0.64) | 0.79 | (0.72, 0.86) |
| | 3-4 days | | | 0.52 | (0.47, 0.58) | 0.69 | (0.61, 0.76) |
| | 5-6 days | | | 0.49 | (0.44, 0.55) | 0.65 | (0.58, 0.73) |
| | Every day | | | 0.45 | (0.41, 0.5) | 0.55 | (0.5, 0.62) |
| Age group at baseline | Early adulthood (16-29) | | | | | 1.00 | - |
| | Early mid-adulthood (30-49) | | | | | 1.14 | (1.02, 1.28) |
| | Late mid-adulthood (50-64) | | | | | 1.34 | (1.2, 1.51) |

| | | Model PA1a | | Model PA2a | | Model PA3a | |
|---|---|---|---|---|---|---|---|
| | **Physical inactivity** | **Odds Ratio** | **95% CI** | **Odds Ratio** | **95% CI** | **Odds Ratio** | **95% CI** |
| | Late adulthood (65+) | | | | | 2.12 | (1.83, 2.45) |
| Sex | Men | | | | | 1.00 | - |
| | women | | | | | 1.67 | (1.56, 1.78) |
| Education | No Qualification | | | | | 1.00 | - |
| | A-Level, GCSE, other qualification | | | | | 0.81 | (0.73, 0.91) |
| | Degree or other higher qualification | | | | | 0.72 | (0.63, 0.81) |
| Ethnicity | White | | | | | 1.00 | - |
| | black | | | | | 1.07 | (0.88, 1.3) |
| | Indian | | | | | 1.25 | (0.96, 1.62) |
| | Pakistani/ Bangladeshi | | | | | 1.71 | (1.35, 2.16) |
| | other Asian/other | | | | | 1.23 | (0.98, 1.55) |
| Occupational Class | Not employed | | | | | 1.00 | - |
| | Management & professional | | | | | 0.99 | (0.81, 1.21) |
| | intermediate | | | | | 0.88 | (0.72, 1.08) |
| | routine | | | | | 1.06 | (0.86, 1.29) |
| Income quintiles | 1 (low) | | | | | 1.00 | - |
| | 2 | | | | | 0.94 | (0.83, 1.05) |
| | 3 | | | | | 0.93 | (0.83, 1.05) |
| | 4 | | | | | 0.84 | (0.74, 0.95) |

| | | Model PA1a | | Model PA2a | | Model PA3a | |
|---|---|---|---|---|---|---|---|
| | Physical inactivity | Odds Ratio | 95% CI | Odds Ratio | 95% CI | Odds Ratio | 95% CI |
| | 5 (high) | | | | | 0.74 | (0.65, 0.83) |
| Working status | Not employed | | | | | 1.00 | - |
| | full-time employed | | | | | 0.86 | (0.71, 1.05) |
| | part-time employed | | | | | 0.89 | (0.74, 1.08) |
| Number of children in household | 0 | | | | | 1.00 | - |
| | 1 | | | | | 0.96 | (0.85, 1.07) |
| | 2 | | | | | 0.79 | (0.69, 0.9) |
| | 3 or more | | | | | 1.03 | (0.83, 1.28) |
| Cohabiting at baseline | Single, divorced, widowed | | | | | 1.00 | - |
| | married or cohabiting | | | | | 1.00 | (0.9, 1.11) |
| Household size | 1 | | | | | 1.00 | - |
| | 2 | | | | | 0.87 | (0.76, 0.99) |
| | 3-4 | | | | | 0.88 | (0.76, 1.01) |
| | 5 or more | | | | | 0.76 | (0.63, 0.91) |
| | GHQ At baseline | | | | | 1.02 | (1.01, 1.02) |
| Self-rated general health | Good or excellent | | | | | 1.00 | - |
| | fair or poor | | | | | 1.37 | (1.22, 1.55) |
| | sf12_base | | | | | 0.98 | (0.97, 0.98) |
| Wave outcome observed | UKHLS 7 | | | | | 1.00 | - |
| | UKHLS 9 | | | | | 0.91 | (0.76, 1.08) |
| | UKHLS 11 | | | | | 1.08 | (0.91, 1.29) |

| | Model PA1a | | Model PA2a | | Model PA3a | |
|---|---|---|---|---|---|---|
| Physical inactivity | Odds Ratio | 95% CI | Odds Ratio | 95% CI | Odds Ratio | 95% CI |
| UKHLS 13 | | | | | 0.99 | (0.87, 1.13) |

**Table S 6:** Regression results for LCA for fruit and vegetable consumption; linear regression models predicting average daily fruit and vegetable intake across latent caregiving intensity classes among UKHLS participants (n=25,049), showing pooled coefficient estimates from multiple imputation (m=10) and accounting for complex survey design and household-level clustering. Results are shown for three models: DIET1a (unadjusted), DIET2a (adjusted for fruit and vegetable intake at baseline), and DIET3a (adjusted for selected covariates).

| | | **Model DIET1a** | | **Model DIET2a** | | **Model DIET3a** | |
|---|---|---|---|---|---|---|---|
| | **Fruit and vegetable consumption** | **Coeff.** | **95% CI** | **Coeff.** | **95% CI** | **Coeff.** | **95% CI** |
| Latent Class | No care | Ref. | - | Ref. | - | Ref. | - |
| | Temporary | 0.1 | (0, 0.2) | 0.0 | (-0.1, 0.2) | 0.1 | (-0.1, 0.2) |
| | Former long | 0.2 | (0.1, 0.4) | 0.1 | (-0.1, 0.2) | 0.1 | (-0.1, 0.2) |
| | Recurrent | 0.2 | (0, 0.4) | 0.1 | (-0.1, 0.3) | 0.1 | (-0.1, 0.3) |
| | Emerging-short | 0.2 | (0.1, 0.4) | 0.2 | (0, 0.3) | 0.1 | (0, 0.2) |
| | Former-short | 0.2 | (0.1, 0.4) | 0.1 | (0, 0.2) | 0.1 | (0, 0.2) |
| | Long-term | 0.1 | (-0.1, 0.2) | 0.0 | (-0.1, 0.1) | 0.0 | (-0.1, 0.1) |
| | Emerging long | 0.1 | (-0.1, 0.2) | 0.0 | (-0.1, 0.2) | 0.0 | (-0.2, 0.1) |
| Portions fruit / vegetable at baseline | 0 | | | Ref. | - | Ref. | - |
| | 1-3 | | | 2.0 | (1.8, 2.3) | 1.7 | (1.5, 1.9) |
| | 4 | | | 3.2 | (2.9, 3.4) | 2.7 | (2.5, 2.9) |
| | 5 or more | | | 4.0 | (3.8, 4.2) | 3.4 | (3.2, 3.7) |
| Age group at baseline | Early adulthood (16-29) | | | | | Ref. | - |
| | Early mid-adulthood (30-49) | | | | | 0.1 | (0, 0.2) |
| | Late mid-adulthood (50-64) | | | | | 0.3 | (0.2, 0.4) |
| | Late adulthood (65+) | | | | | 0.2 | (0, 0.3) |

| | | Model DIET1a | | Model DIET2a | | Model DIET3a | |
|---|---|---|---|---|---|---|---|
| | **Fruit and vegetable consumption** | **Coeff.** | **95% CI** | **Coeff.** | **95% CI** | **Coeff.** | **95% CI** |
| Sex | Men | | | | | Ref. | - |
| | women | | | | | 0.2 | (0.1, 0.2) |
| Education | No Qualification | | | | | Ref. | - |
| | A-Level, GCSE, other qualification | | | | | 0.3 | (0.2, 0.4) |
| | Degree or other higher qualification | | | | | 0.7 | (0.6, 0.8) |
| Ethnicity | White | | | | | Ref. | - |
| | black | | | | | 0.0 | (-0.2, 0.2) |
| | Indian | | | | | -0.1 | (-0.3, 0.1) |
| | Pakistani/ Bangladeshi | | | | | -0.5 | (-0.8, -0.3) |
| | other Asian/other | | | | | 0.3 | (0.1, 0.5) |
| Occupational Class | Not employed | | | | | Ref. | - |
| | Management & professional | | | | | -0.3 | (-0.5, -0.1) |
| | intermediate | | | | | -0.3 | (-0.5, -0.1) |
| | routine | | | | | -0.5 | (-0.7, -0.3) |
| Income quintiles | 1 (low) | | | | | Ref. | - |
| | 2 | | | | | 0.0 | (-0.1, 0.1) |
| | 3 | | | | | 0.1 | (0, 0.2) |
| | 4 | | | | | 0.2 | (0.1, 0.3) |
| | 5 (high) | | | | | 0.4 | (0.3, 0.5) |
| Working status | Not employed | | | | | Ref. | - |

| | | Model DIET1a | | Model DIET2a | | Model DIET3a | |
|---|---|---|---|---|---|---|---|
| | **Fruit and vegetable consumption** | **Coeff.** | **95% CI** | **Coeff.** | **95% CI** | **Coeff.** | **95% CI** |
| | full-time employed | | | | | 0.3 | (0.1, 0.4) |
| | part-time employed | | | | | 0.4 | (0.2, 0.6) |
| Number of children in household | 0 | | | | | Ref. | - |
| | 1 | | | | | -0.1 | (-0.2, 0) |
| | 2 | | | | | 0.0 | (-0.1, 0.1) |
| | 3 or more | | | | | -0.2 | (-0.4, 0) |
| Cohabiting at baseline | Single, divorced, widowed | | | | | Ref. | - |
| | married or cohabiting | | | | | 0.2 | (0.1, 0.3) |
| Number of people living in the household | 1 | | | | | Ref. | - |
| | 2 | | | | | 0.0 | (-0.2, 0.1) |
| | 3-4 | | | | | 0.0 | (-0.1, 0.1) |
| | 5 or more | | | | | 0.0 | (-0.1, 0.2) |
| | GHQ At baseline | | | | | 0.0 | (0, 0) |
| Self-rated general health | Good or excellent | | | | | Ref. | - |
| | fair or poor | | | | | -0.2 | (-0.3, -0.1) |
| Wave outcome observed | UKHLS 7 | | | | | Ref. | - |
| | UKHLS 9 | | | | | 0.0 | (-0.2, 0.1) |
| | UKHLS 11 | | | | | 0.0 | (-0.1, 0.2) |
| | UKHLS 13 | | | | | 0.1 | (0, 0.2) |

**Table S.7:** Regression results for LCA for Problematic Drinking**;** logistic regression models predicting problematic drinking across latent caregiving intensity classes among UKHLS participants (n=25,049), showing pooled odds ratio estimates from multiple imputation (m=10) and accounting for complex survey design and household-level clustering. Results are shown for three models: ALC1a (unadjusted), ALC2a (adjusted for drinks frequency at baseline), and ALC3a (adjusted for selected covariates).

| | | **Model ALC1a** | | **Model ALC2a** | | **Model ALC3a** | |
|---|---|---|---|---|---|---|---|
| | **Problematic drinking** | **Odds Ratio** | **95% CI** | **Odds Ratio** | **95% CI** | **Odds Ratio** | **95% CI** |
| Latent Class | No care | 1.00 | - | 1.00 | - | 1.00 | - |
| | Temporary | 0.89 | (0.79, 1) | 0.86 | (0.75, 0.97) | 0.97 | (0.84, 1.11) |
| | Former long | 0.86 | (0.74, 1) | 0.76 | (0.64, 0.9) | 0.90 | (0.75, 1.07) |
| | Recurrent | 0.74 | (0.61, 0.89) | 0.67 | (0.54, 0.83) | 0.75 | (0.59, 0.94) |
| | Emerging-short | 0.92 | (0.81, 1.05) | 0.86 | (0.74, 0.99) | 0.88 | (0.75, 1.03) |
| | Former-short | 0.88 | (0.78, 0.99) | 0.80 | (0.7, 0.91) | 0.95 | (0.83, 1.09) |
| | Long-term | 0.82 | (0.72, 0.93) | 0.83 | (0.72, 0.96) | 0.87 | (0.74, 1.01) |
| | Emerging long | 0.92 | (0.8, 1.07) | 0.90 | (0.76, 1.05) | 0.91 | (0.77, 1.08) |
| Walking at baseline | Non-drinker | | | 1.00 | - | 1.00 | - |
| | Monthly/weekly | | | 5.08 | (4.16, 6.21) | 3.75 | (3.04, 4.61) |
| | 1-4 days/week | | | 24.71 | (20.3, 30.09) | 23.10 | (18.8, 28.37) |
| | 5+ days a week | | | 76.17 | (61.02, 95.07) | 97.86 | (77.17, 124.1) |
| Age group at baseline | Early adulthood (16-29) | | | | | 1.00 | - |
| | Early mid-adulthood (30-49) | | | | | 0.69 | (0.6, 0.78) |
| | Late mid-adulthood (50-64) | | | | | 0.43 | (0.38, 0.49) |
| | Late adulthood (65+) | | | | | 0.18 | (0.15, 0.21) |

| | | Model ALC1a | | Model ALC2a | | Model ALC3a | |
|---|---|---|---|---|---|---|---|
| | **Problematic drinking** | **Odds Ratio** | **95% CI** | **Odds Ratio** | **95% CI** | **Odds Ratio** | **95% CI** |
| Sex | Men | | | | | 1.00 | - |
| | women | | | | | 1.55 | (1.44, 1.67) |
| Education | No Qualification | | | | | 1.00 | - |
| | A-Level, GCSE, other qualification | | | | | 1.12 | (0.98, 1.28) |
| | Degree or other higher qualification | | | | | 1.06 | (0.92, 1.23) |
| Ethnicity | White | | | | | 1.00 | - |
| | black | | | | | 0.60 | (0.46, 0.77) |
| | Indian | | | | | 0.39 | (0.29, 0.54) |
| | Pakistani/ Bangladeshi | | | | | 0.19 | (0.1, 0.36) |
| | other Asian/other | | | | | 0.63 | (0.47, 0.85) |
| Occupational Class | Not employed | | | | | 1.00 | - |
| | Management & professional | | | | | 1.02 | (0.8, 1.29) |
| | intermediate | | | | | 0.91 | (0.72, 1.15) |
| | routine | | | | | 1.00 | (0.79, 1.26) |
| Income quintiles | 1 (low) | | | | | 1.00 | - |
| | 2 | | | | | 1.12 | (0.98, 1.29) |
| | 3 | | | | | 1.28 | (1.12, 1.47) |
| | 4 | | | | | 1.42 | (1.24, 1.63) |
| | 5 (high) | | | | | 1.61 | (1.4, 1.86) |
| Working status | Not employed | | | | | 1.00 | - |

| | | Model ALC1a | | Model ALC2a | | Model ALC3a | |
|---|---|---|---|---|---|---|---|
| | **Problematic drinking** | **Odds Ratio** | **95% CI** | **Odds Ratio** | **95% CI** | **Odds Ratio** | **95% CI** |
| | full-time employed | | | | | 0.87 | (0.69, 1.1) |
| | part-time employed | | | | | 1.04 | (0.83, 1.3) |
| Number of children in household | 0 | | | | | 1.00 | - |
| | 1 | | | | | 1.24 | (1.09, 1.41) |
| | 2 | | | | | 1.48 | (1.28, 1.72) |
| | 3 or more | | | | | 1.01 | (0.78, 1.32) |
| Cohabiting at baseline | Single, divorced, widowed | | | | | 1.00 | - |
| | married or cohabiting | | | | | 1.04 | (0.93, 1.17) |
| | 1 | | | | | 1.00 | - |
| | 2 | | | | | 1.16 | (1, 1.35) |
| | 3-4 | | | | | 1.20 | (1.02, 1.41) |
| | 5 or more | | | | | 1.23 | (0.99, 1.54) |
| | GHQ At baseline | | | | | 1.00 | (1, 1.01) |
| Self-rated general health | Good or excellent | | | | | 1.00 | - |
| | fair or poor | | | | | 0.74 | (0.66, 0.83) |
| Wave outcome observed | UKHLS 7 | | | | | 1.00 | - |
| | UKHLS 9 | | | | | 0.92 | (0.73, 1.15) |
| | UKHLS 11 | | | | | 0.66 | (0.54, 0.8) |
| | UKHLS 13 | | | | | 0.59 | (0.5, 0.68) |

**Table S.8:** Regression results for LCA for Smoking; logistic regression models predicting smoking status across latent caregiving intensity classes among UKHLS participants (n=25,049), showing pooled odds ratio estimates from multiple imputation (m=10) and accounting for survey weights and household-level clustering. Results are shown for three models: SMOK1a (unadjusted), SMOK2a (adjusted for smoking status at baseline), and SMOK3a (adjusted for selected covariates).

| | | Model SMOK1a | | Model SMOK2a | | Model SMOK3a | |
|---|---|---|---|---|---|---|---|
| | | Odds Ratio | 95% CI | Odds Ratio | 95% CI | Odds Ratio | 95% CI |
| Latent Class | No care | 1.00 | - | 1.00 | - | 1.00 | - |
| | Temporary | 1.10 | (0.92, 1.3) | 1.08 | (0.87, 1.34) | 1.12 | (0.89, 1.41) |
| | Former long | 1.05 | (0.84, 1.31) | 1.12 | (0.84, 1.5) | 1.31 | (0.96, 1.8) |
| | Recurrent | 1.58 | (1.22, 2.04) | 1.50 | (1.06, 2.12) | 1.67 | (1.17, 2.4) |
| | Emerging-short | 0.94 | (0.76, 1.16) | 0.89 | (0.69, 1.17) | 0.99 | (0.74, 1.32) |
| | Former-short | 1.00 | (0.84, 1.2) | 0.98 | (0.79, 1.22) | 1.04 | (0.83, 1.31) |
| | Long-term | 1.18 | (0.98, 1.43) | 1.09 | (0.86, 1.37) | 1.05 | (0.83, 1.34) |
| | Emerging long | 1.15 | (0.93, 1.44) | 1.04 | (0.8, 1.36) | 1.08 | (0.81, 1.43) |
| Smoking at baseline | Non-smoker | | | 1.00 | - | 1.00 | - |
| | Ex-smoker | | | 3.41 | (2.67, 4.34) | 4.15 | (3.26, 5.29) |
| | Current smoker | | | 96.01 | (77.03, 119.66) | 92.52 | (74.29, 115.22) |
| Age group at baseline | Early adulthood (16-29) | | | | | 1.00 | - |
| | Early mid-adulthood (30-49) | | | | | 0.77 | (0.63, 0.94) |
| | Late mid-adulthood (50-64) | | | | | 0.70 | (0.57, 0.87) |
| | Late adulthood (65+) | | | | | 0.28 | (0.21, 0.37) |
| Sex | Men | | | | | 1.00 | - |
| | women | | | | | 0.88 | (0.78, 1) |

| | | Model SMOK1a | | Model SMOK2a | | Model SMOK3a | |
|---|---|---|---|---|---|---|---|
| | | Odds Ratio | 95% CI | Odds Ratio | 95% CI | Odds Ratio | 95% CI |
| Education | No Qualification | | | | | 1.00 | - |
| | A-Level, GCSE, other qualification | | | | | 0.76 | (0.64, 0.92) |
| | Degree or other higher qualification | | | | | 0.54 | (0.43, 0.67) |
| Ethnicity | White | | | | | 1.00 | - |
| | black | | | | | 1.29 | (0.87, 1.93) |
| | Indian | | | | | 0.65 | (0.39, 1.08) |
| | Pakistani/ Bangladeshi | | | | | 0.98 | (0.64, 1.5) |
| | other Asian/other | | | | | 1.90 | (1.16, 3.13) |
| Occupational Class | Not employed | | | | | 1.00 | - |
| | Management & professional | | | | | 0.80 | (0.51, 1.27) |
| | intermediate | | | | | 0.83 | (0.53, 1.31) |
| | routine | | | | | 0.99 | (0.64, 1.54) |
| Income quintiles | 1 (low) | | | | | 1.00 | - |
| | 2 | | | | | 0.91 | (0.75, 1.1) |
| | 3 | | | | | 0.94 | (0.77, 1.16) |
| | 4 | | | | | 0.81 | (0.65, 1) |
| | 5 (high) | | | | | 0.64 | (0.51, 0.81) |
| Working status | Not employed | | | | | 1.00 | - |
| | full-time employed | | | | | 0.82 | (0.52, 1.29) |

| | | Model SMOK1a | | Model SMOK2a | | Model SMOK3a | |
|---|---|---|---|---|---|---|---|
| | | **Odds Ratio** | **95% CI** | **Odds Ratio** | **95% CI** | **Odds Ratio** | **95% CI** |
| | part-time employed | | | | | 0.82 | (0.53, 1.27) |
| Number of children in household | 0 | | | | | 1.00 | - |
| | 1 | | | | | 1.09 | (0.88, 1.34) |
| | 2 | | | | | 0.91 | (0.72, 1.16) |
| | 3 or more | | | | | 0.62 | (0.42, 0.93) |
| Cohabiting at baseline | Single, divorced, widowed | | | | | 1.00 | - |
| | married or cohabiting | | | | | 0.74 | (0.62, 0.88) |
| | 1 | | | | | 1.00 | - |
| | 2 | | | | | 0.91 | (0.73, 1.13) |
| | 3-4 | | | | | 1.13 | (0.89, 1.44) |
| | 5 or more | | | | | 1.57 | (1.12, 2.21) |
| | GHQ At baseline | | | | | 1.00 | (0.99, 1.01) |
| Self-rated general health | Good or excellent | | | | | 1.00 | - |
| | fair or poor | | | | | 1.05 | (0.89, 1.24) |
| Wave outcome observed | UKHLS 7 | | | | | 1.00 | - |
| | UKHLS 9 | | | | | 0.83 | (0.62, 1.1) |
| | UKHLS 11 | | | | | 0.69 | (0.52, 0.9) |
| | UKHLS 13 | | | | | 0.45 | (0.37, 0.55) |

### S9: Interactions

**Table S.9:** Wald-test p-values for interaction terms between latent caregiving intensity classes and Observed Transition groups, predicting health behaviours among UKHLS participants (n=25,049). Estimates account for complex survey design, clustering at the household level, and multiple imputation (m=10).

| | **Latent class variable** | | **observed typology** | |
|---|---|---|---|---|
| | Sex | Age groups | Sex | Age groups |
| Physical inactivity | 0.05 | 0.71 | 0.30 | 0.79 |
| Fruit and vegetable consumption | 0.74 | 0.01 | 0.68 | 0.02 |
| Problematic drinking | 0.88 | 0.44 | 0.71 | 0.06 |
| Smoking | 0.34 | 0.59 | 0.02 | 0.88 |

**Table S.10:** Regression results for LCA for physical inactivity, stratified by sex. Logistic regression models predicting physical inactivity across latent caregiving intensity classes, stratified by sex, among UKHLS participants (n=25,049), showing pooled odds ratios from multiple imputation (m=10) and accounting for complex survey design and household-level clustering. Estimates are from the fully adjusted model (PA3a).

| | | **Men** | | **Women** | |
|---|---|---|---|---|---|
| | **Physical inactivity** | **Odds Ratio** | **95% CI** | **Odds Ratio** | **95% CI** |
| Latent Class | No care | 1.00 | - | 1.00 | - |
| | Temporary | 0.85 | (0.71, 1.03) | 0.96 | (0.82, 1.12) |
| | Former-long | 1.44 | (1.10, 1.87) | 0.96 | (0.79, 1.16) |
| | Recurrent | 0.65 | (0.46, 0.94) | 0.65 | (0.50, 0.85) |
| | Emerging-short | 1.00 | (0.81, 1.23) | 0.98 | (0.82, 1.17) |
| | Former-short | 0.90 | (0.75, 1.08) | 0.98 | (0.84, 1.15) |
| | Long-term | 1.14 | (0.89, 1.44) | 0.83 | (0.70, 0.98) |
| | Emerging-long | 1.01 | (0.78, 1.30) | 0.89 | (0.73, 1.09) |

*Adjusted for walking frequency at baseline, age group, education, ethnicity, occupational class, household income quintile, working status, number of children in the household, cohabitation status, household size, GHQ-12 score, self-rated general health, SF-12 PCS and wave of outcome observation.

**Table S.11:** Regression results for LCA for fruit and vegetable consumption, stratified by age group. Linear regression models predicting average daily fruit and vegetable intake across latent caregiving intensity classes, stratified by age group, among UKHLS participants (n=25,049), showing pooled coefficient estimates from multiple imputation (m=10) and accounting for complex survey design and household-level clustering. Estimates are from the fully adjusted model (DIET3a).

| | | 16-29 | | 30-49 | | 50-64 | | 65+ | |
|---|---|---|---|---|---|---|---|---|---|
| | **Fruit and vegetable consumption** | **Coeff.*** | **95% CI** | **Coeff.*** | **95% CI** | **Coeff.*** | **95% CI** | **Coeff.*** | **95% CI** |
| Latent Class | No care | Ref. | - | Ref. | - | Ref. | - | Ref. | - |
| | Temporary | -0.04 | (-0.39, 0.31) | 0.07 | (-0.14, 0.27) | 0.11 | (-0.09, 0.31) | 0.03 | (-0.22, 0.29) |
| | Former-long | 0.07 | (-0.56, 0.69) | -0.20 | (-0.46, 0.06) | 0.13 | (-0.09, 0.35) | 0.31 | (0.04, 0.58) |
| | Recurrent | -0.08 | (-0.81, 0.64) | -0.25 | (-0.54, 0.05) | 0.29 | (-0.01, 0.58) | 0.52 | (0.06, 0.98) |
| | Emerging-short | 0.00 | (-0.36, 0.35) | 0.02 | (-0.16, 0.21) | 0.27 | (0.03, 0.52) | 0.07 | (-0.26, 0.40) |
| | Former-short | -0.09 | (-0.43, 0.25) | 0.04 | (-0.16, 0.25) | 0.19 | (0.00, 0.37) | 0.14 | (-0.09, 0.37) |
| | Long-term | 0.02 | (-0.51, 0.55) | -0.05 | (-0.22, 0.12) | 0.23 | (0.01, 0.45) | -0.16 | (-0.47, 0.14) |
| | Emerging-long | -0.55 | (-0.92, -0.18) | 0.02 | (-0.22, 0.26) | 0.16 | (-0.08, 0.40) | -0.02 | (-0.39, 0.35) |

*Adjusted for fruit and vegetable intake at baseline, sex, education, ethnicity, occupational class, household income quintile, working status, number of children in the household, cohabitation status, household size, GHQ-12 score, self-rated general health, and wave of outcome observation.

### S10: Observed transitions

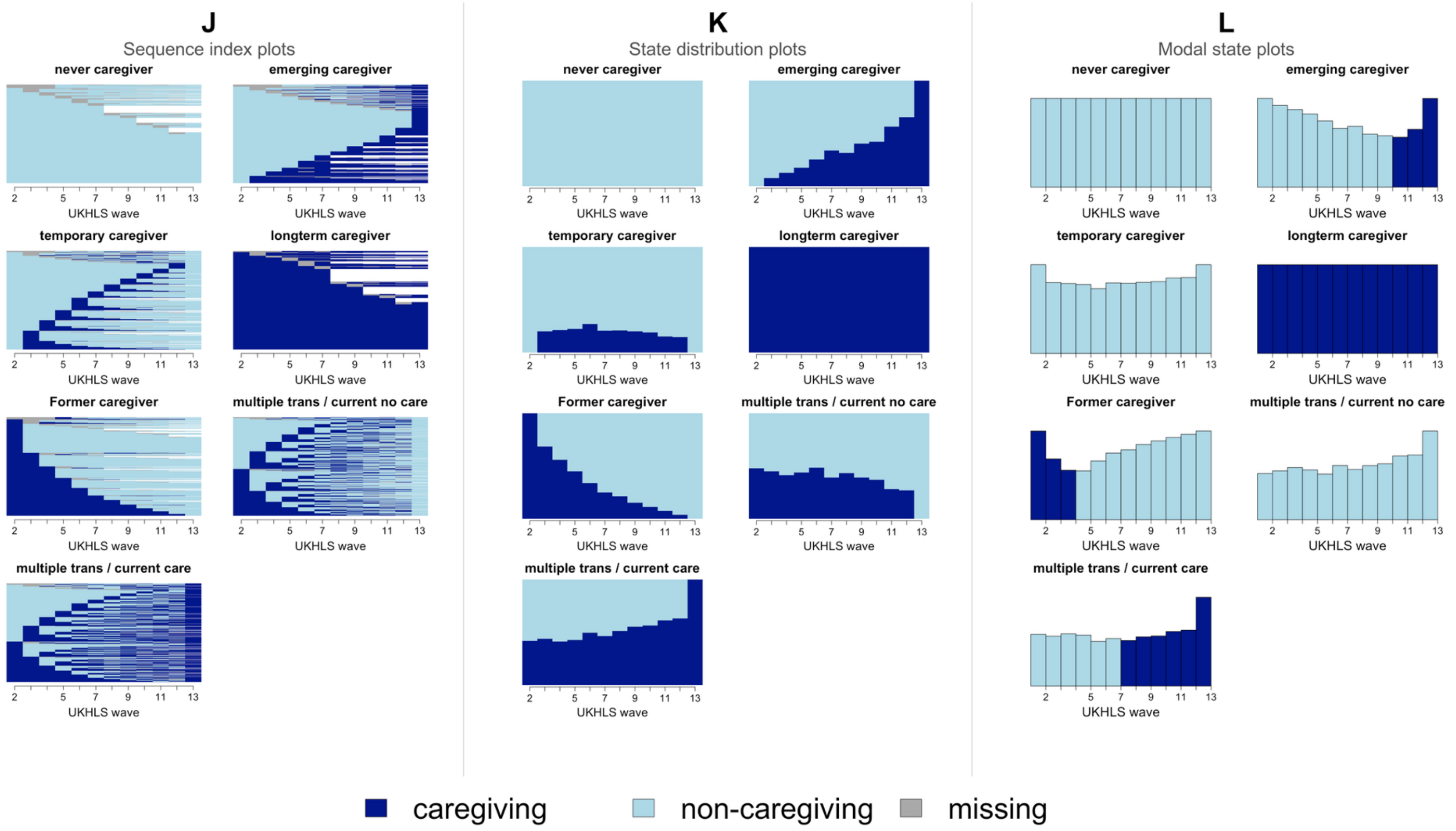

**Figure S.9:** Caregiving trajectories by observed transition typology, UK Household Longitudinal Study waves 2 to 13 (n = 25,049).
(J) Sequence index plots, (K) state distribution plots and (L) modal state plots for each of the seven observed transition groups. Dark blue denotes caregiving, light blue non-caregiving and grey missing data**.**

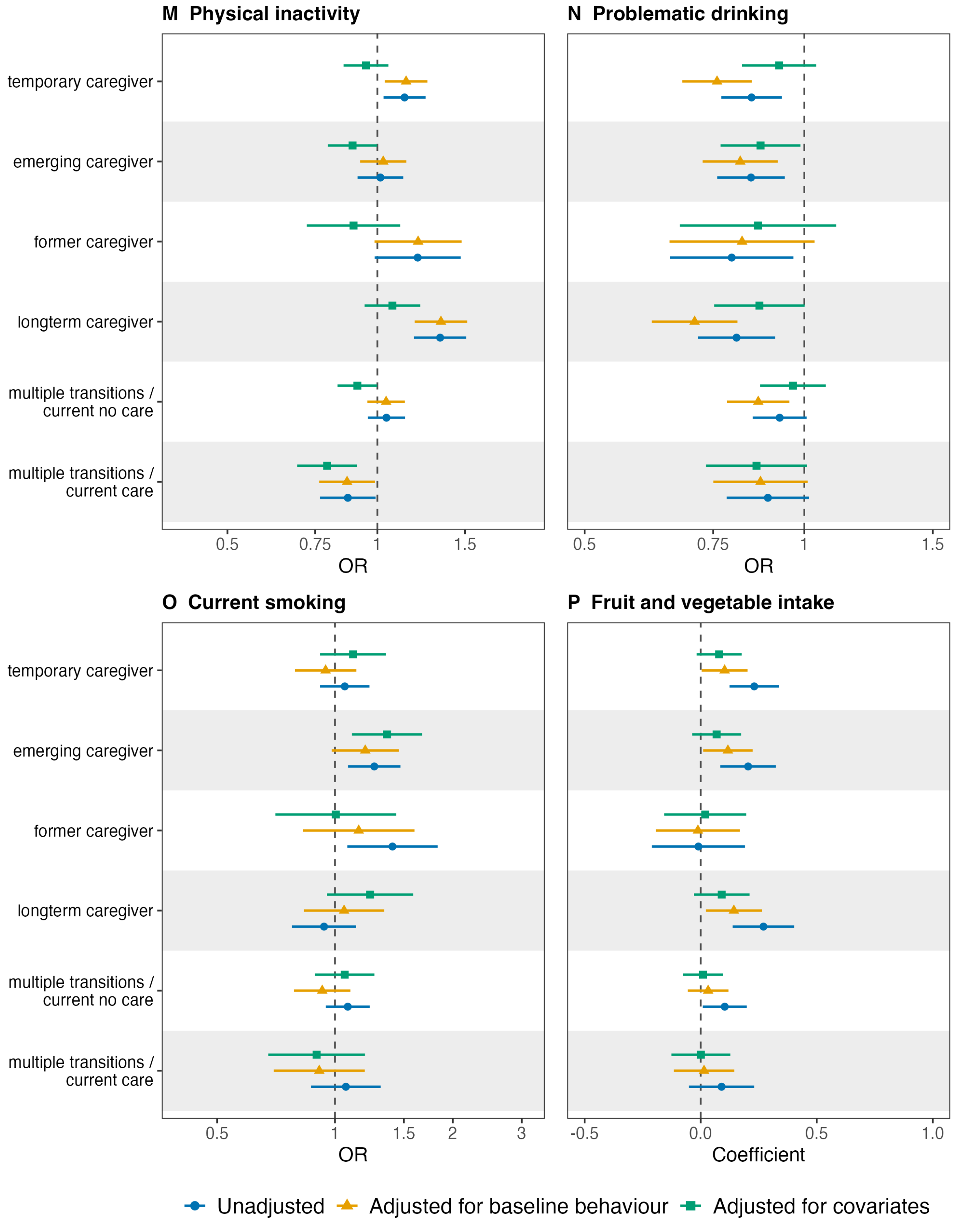

**Figure S.10:** Association between observed transition typology and health behaviours, UK Household Longitudinal Study (n = 25,049). Panels show (M) physical inactivity, (N) problematic drinking, (O) current smoking and (P) average daily fruit and vegetable consumption. Points are odds ratios (M, N, O) or linear regression coefficients (P) with 95% confidence intervals, pooled across ten imputed datasets and accounting for the complex survey design and household-level clustering. Never caregiver is the reference group. Estimates are shown unadjusted, adjusted for the corresponding baseline health behaviour, and adjusted for covariates.

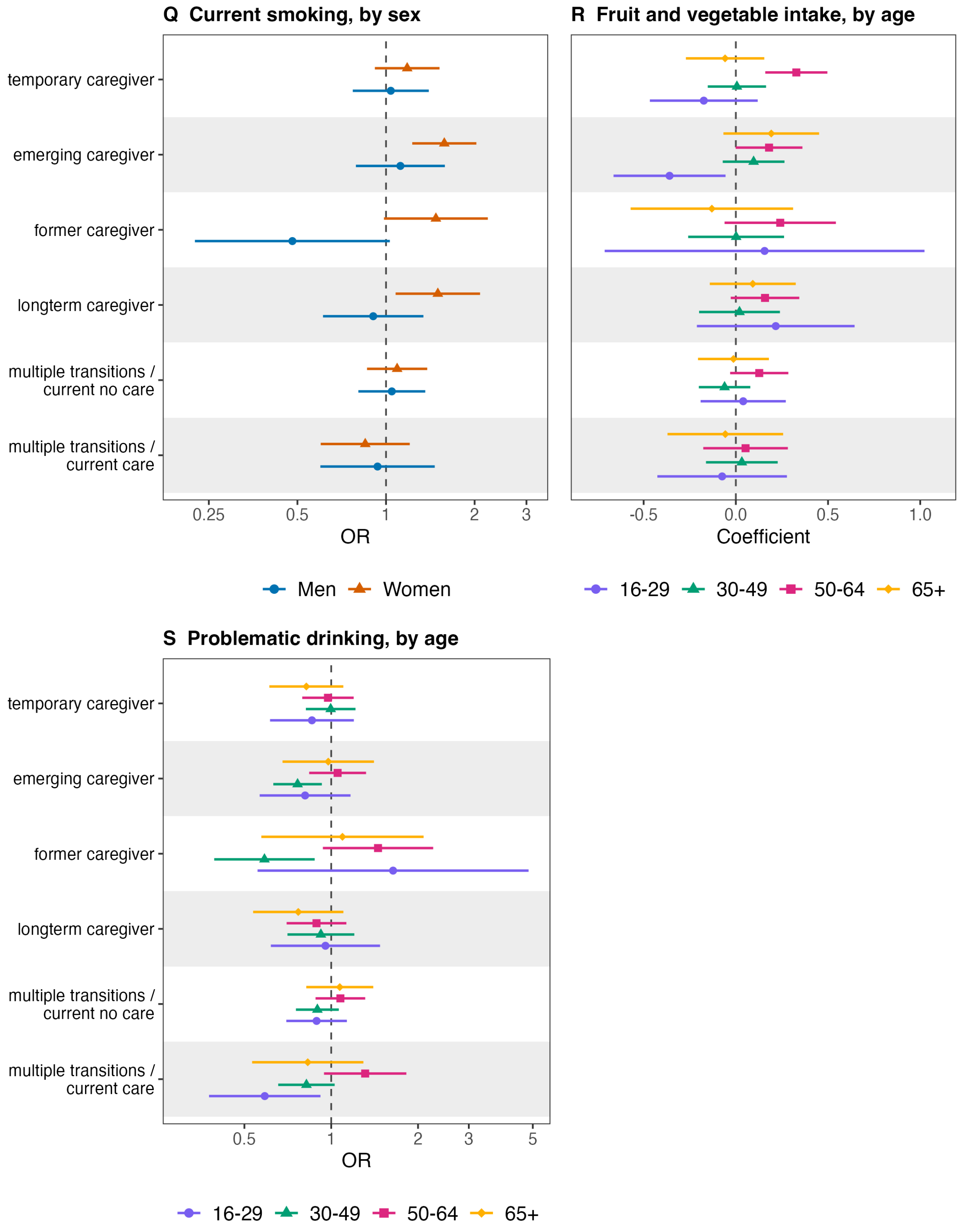

**Figure S.11:** Association between observed transition typology and health behaviours, stratified by sex and age group, UK Household Longitudinal Study (n = 25,049). Panels show (Q) current smoking by sex, (R) average daily fruit and vegetable consumption by age group and (S) problematic drinking by age group. Points are odds ratios (Q, S) or linear regression coefficients (R) with 95% confidence intervals from fully adjusted models, pooled across ten imputed datasets and accounting for the complex survey design and household-level clustering. Never caregiver is the reference group. Adjusted for fruit and vegetable intake at baseline, sex, education, ethnicity, occupational class, household income quintile, working status, number of children in the household, cohabitation status, household size, GHQ-12 score, self-rated general health, and wave of outcome observation.

### S11: Sequence Analysis

**Sequence analysis (SA) Methods**

**Process**

An alternative approach to LCA is Sequence Analysis (SA) in which categorical time-series variables can be studied  as states or events over time in view of their patterns, transitions, and similarities.[1] While many scholars have argued that LCA is a superior approach compared to SA[2–4], the advantage of sequence analysis (SA) lies in its ability to perform sequence imputation on gaps within a sequence. This approach was developed by Halpin[5] in 2016 and advanced with the release of a new R package by Emery in March 2024[6]. It must be noted that sequence imputation is a fairly new approach that is still in the process of being refined. Besides, it remains an open problem how to perform cluster analyses on imputed data sets[6] but the proposed approach by Halpin[7] was performed in which cluster analysis is performed on the stacked imputed dataset. Nevertheless, Sequence imputation is superior to ‘regular’ multiple imputation for categorical time-series data because it preserves the temporal and sequential structure of the data. In multiple imputation, each time point is treated independently whereas sequence imputation considers the dependency between consecutive time points.[5,7]

Unfortunately, it is only possible to perform sequence imputation on the sequence variable of interesting which is caregiving status for this analysis, but it is not possible to impute missing data of covariate simultaneously within the same package. However, it is possible to run a sperate multiple imputation using Multiple Imputation by Chained equation (MICE) to impute missing covariates with the mice package in R. Following the imputations that occurred, the mice data set and the sequence imputed data set can be merged and pooled regression be performed. The macro flowchart below explains the process.

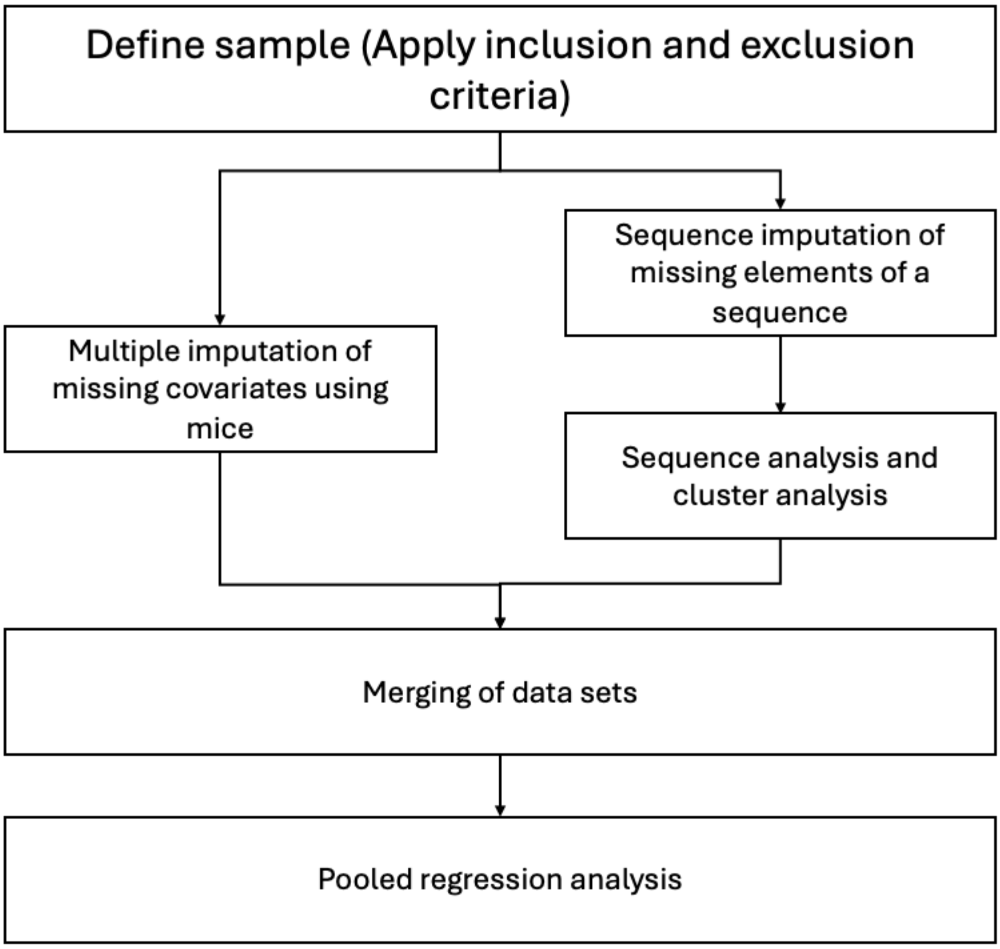

**Figure S.12**: **Overview of the sequence imputation and analysis approach.** Missing covariates were imputed by chained equations and missing sequence states by sequence imputation; the two imputed datasets were merged before pooled regression analysis.

The following steps will be needed to perform SA and subsequent regression on the cluster variable:

***a) Define sample***

After inclusion and exclusion criteria are applied, the patterns of missingness will be assessed for this sample.

***b) MICE***

Multiple imputation by chained equation will be performed using the mice package in R. For this, five imputations will be conducted because missingness of covariates is 17.4% and there are over 20,000 participants over 12 time points in the data set. It was considered that imputing five data set would strike a balance between enhancing accuracy and making the analysis computational feasible. It was decided to perform an imputation with five data sets because literature suggest that at least five imputation are required to handle uncertainty associated with missing data.[8] Also, imputing sequences is computationally intensive, and five imputations is still feasible while providing variation in the estimates. It is also the standard approach that was proposed by Halpin.[7] Therefore, it was considered that five imputations provide a reasonable balance between accuracy and practicality. All covariates that serve for the final regression model were used for the imputation model because all were associated with missingness.

##### c) *Sequence imputation*

Sequence imputation will be performed with the seqimpute package from R. For this, five imputations will be conducted because the number of imputations had to align with the number of imputation from step b (mice). Because the data set was large and due to a high number of distinct sequences, the data was aggregated using the R package WeighedCluster. Then sequence analysis is performed on the aggregated datasets with weights.

##### d) *Dissimilarity measures*

To measure dissimilarity between sequences, a wide range of approaches is available as summarised in the table below. To answer the research question, two approaches were considered most suitable, namely Number of matching sub-sequences (NMS) and optimal matching (OM). NMS was considered suitable because multiple transitions might create complex sequences were individuals transitions between caregiving states. Counting the number of matching sub-sequences allows to identify similarity in the complexity of patterns. Likewise, OM seems like a suitable approach that is flexible and allows to measure dissimilarity and temporal alignment of sequences.[9,10] Since caregiving status only had two states (non-caregiving or caregiving), more complex dissimilarity measures such as time-ward-edit edit distance (TWED) would probably only add complexity while not adding much analytical value.

##### e) Number of clusters

The ideal number of clusters was determined by the following indicators Point Biserial Correlation (PBC), Hubert's Gamma (HG), Hubert's Somers D (HGSD), Hubert's C (HC),

Average Silhouette Width (ASW), Calinski-Harabasz Index (CH), Pseudo R2 (R2). Each cluster solution is assigned a value for any of these indicators and higher values indicate better fit with the exception for Hubert's C (HC) for which lower values indicate better fit. which is available in the publication from Studer.[11] For the analysis, a graph will be produced with the WeightedCluster package to assess all of these quality measures simultaneously.

###### f) Cluster linkage

Several linkages of clustering dissimilarity matrix are available including ward's linkage and average linkage. Average linkage calculates the distance between two clusters as the average of the distance between all pairs of points from the two clusters and it a suitable method if outliers are to be expected.[12] In contrast, ward linkage minimises the variance within clusters by merging the pair that results in the smallest increase in total within-cluster variance, making it effective for producing clusters that are roughly equally sized and cohesive.[13,14] Whether ward linkage or average linkage will be used for a particular cluster solution will depend on the fit indicators and whether the emerging clusters are conceptually plausible.

###### *g) Merge data sets*

Both imputation data sets will be merged by unique identifier for each participant (pidp) and cross-tabulation and assessment of duplicates will be performed to ensure this occurs correctly.

###### *h) Regression and pooled results*

Regression analysis will be performed on each imputed data set and each iteration will also be adjusted for clustering at household level and complex survey design using the svyglm package in R.

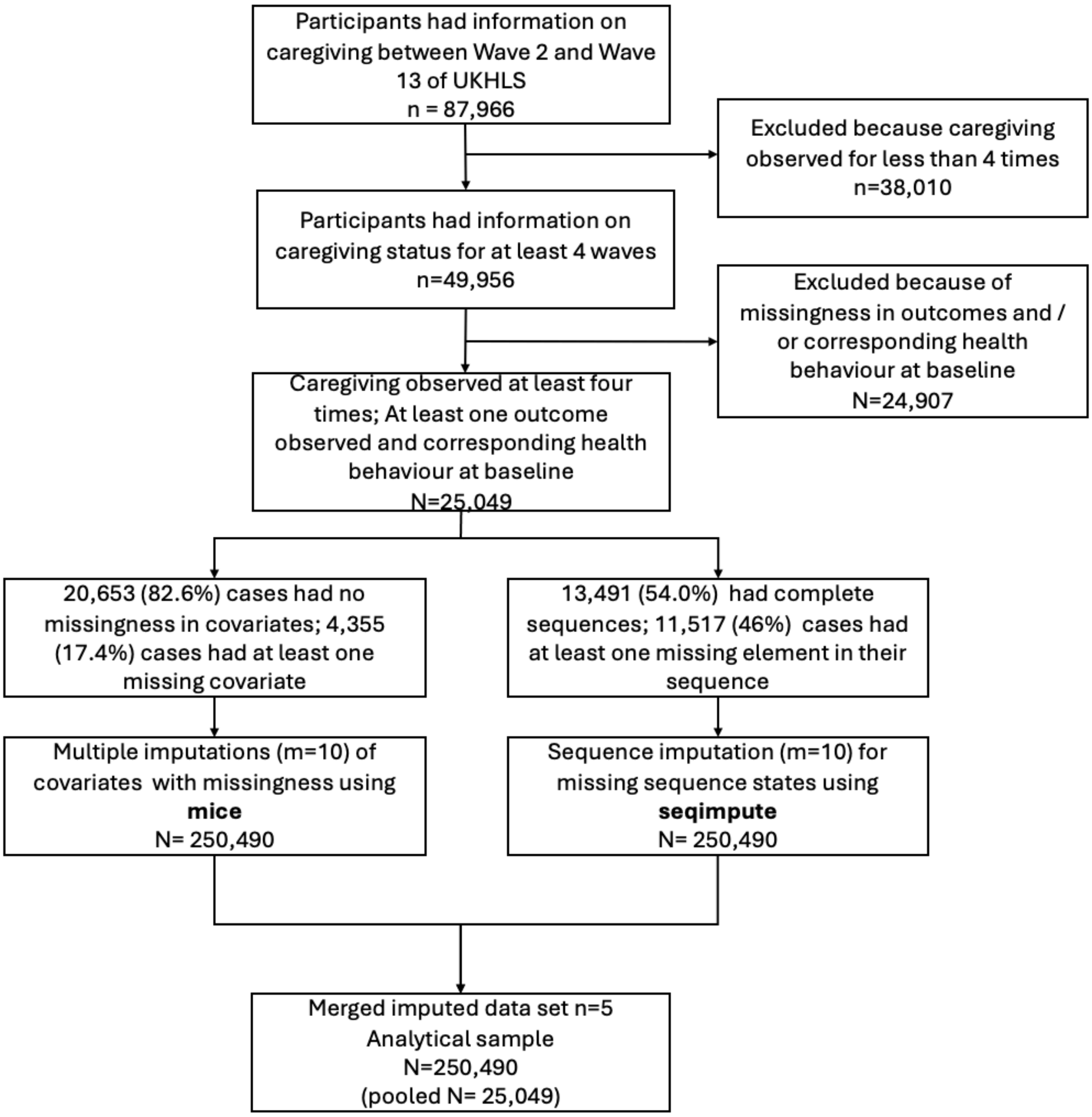

**Figure S.13: Derivation of the analytic sample and imputation structure for the sequence analysis, UK Household Longitudinal Study waves 2 to 13.** Covariate missingness was imputed using mice and missing sequence states using seqimpute, each with ten imputations, giving 250,490 stacked records corresponding to 25,049 participants.

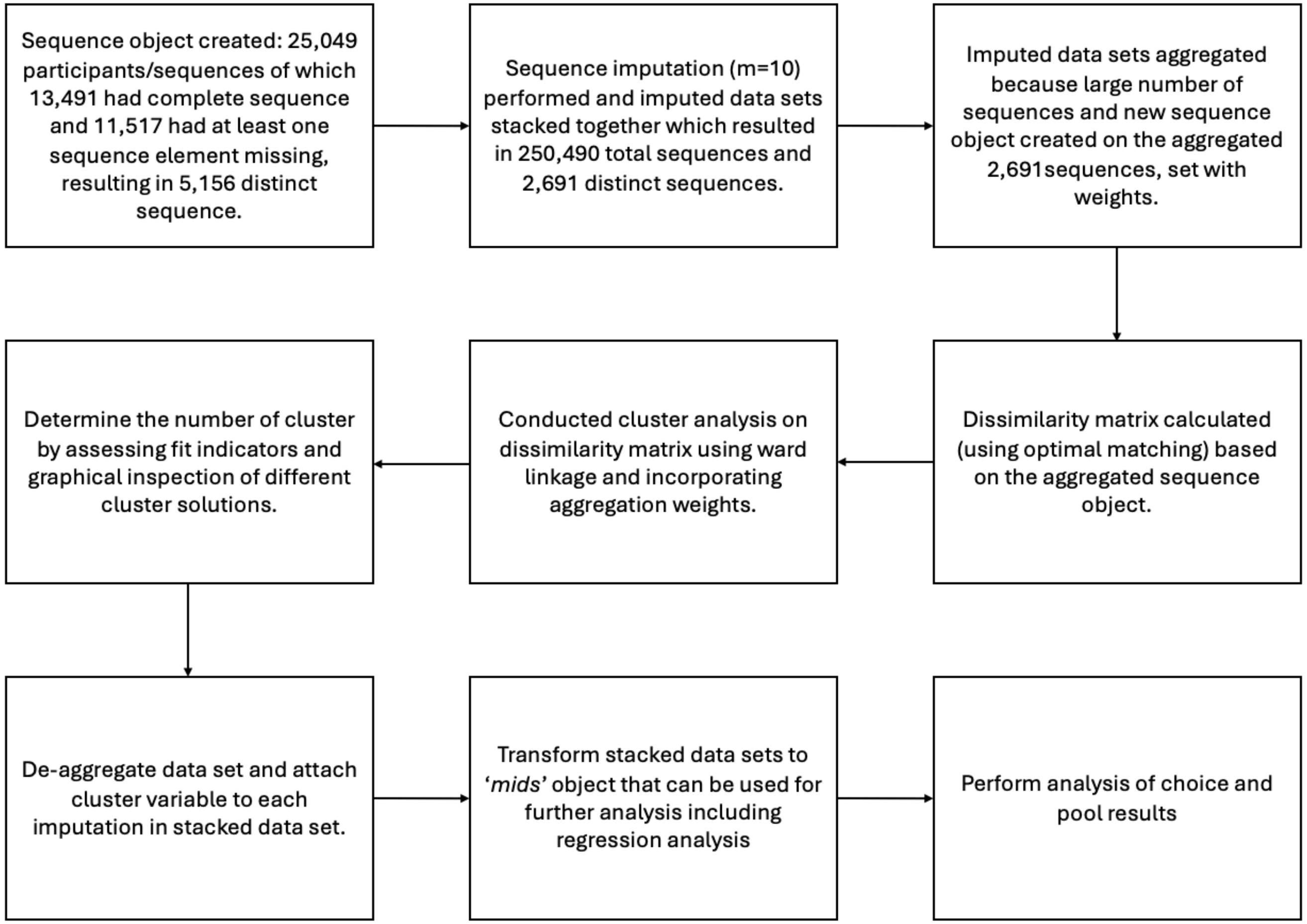

**Figure S.14: Analytical workflow for sequence imputation**, aggregation, clustering and pooled regression. Sequences were aggregated with weights before dissimilarity matrices were computed by optimal matching and clustered using Ward linkage.

#### Sequence analysis

##### Patterns of missingness

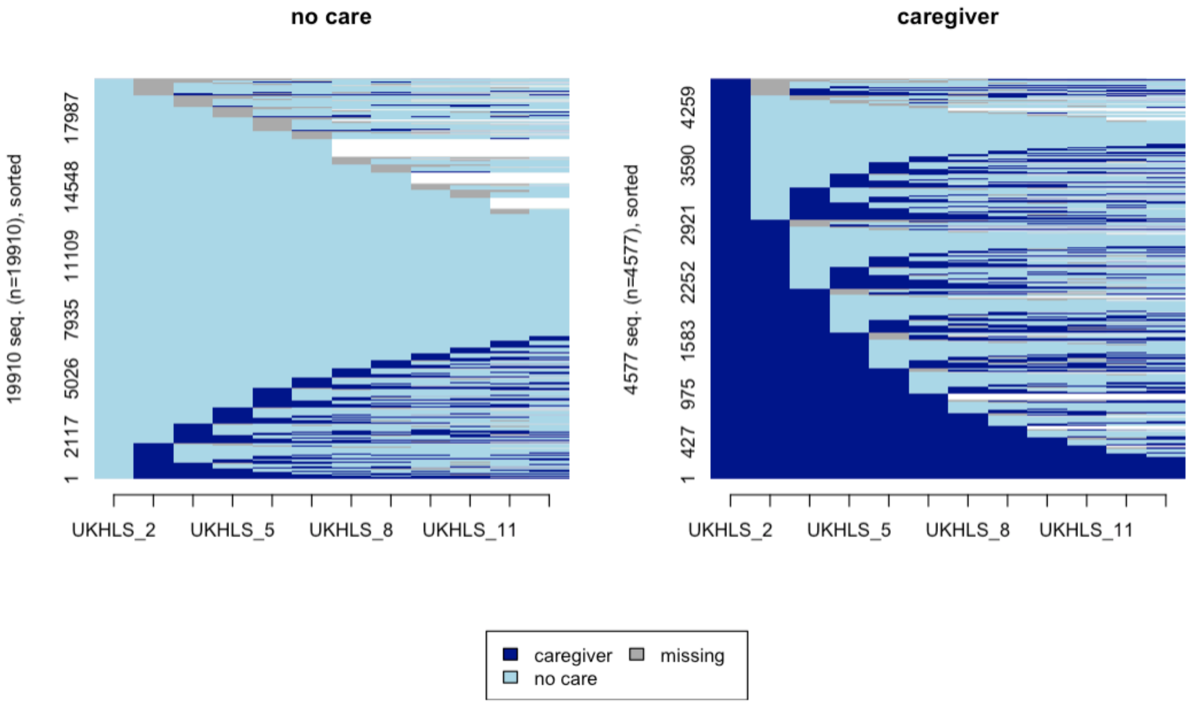

**Figure S. 15: Sequence index plot by caregiving status at baseline,** UK Household Longitudinal Study waves 2 to 13 (n = 25,049). Based on observed data before sequence imputation.

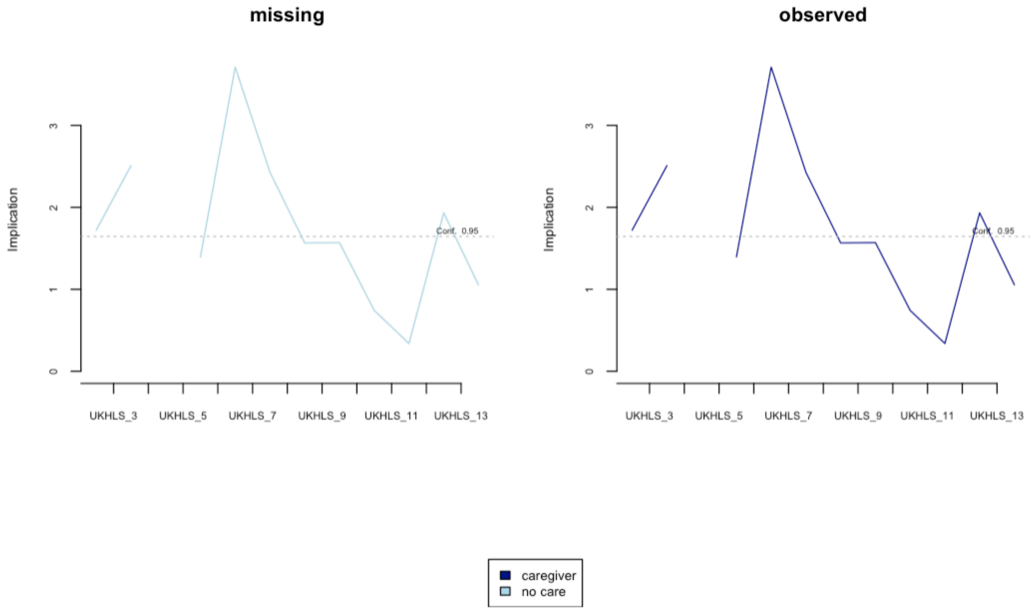

**Figure S.16: Implication statistic for caregiving status by missing and observed group, UK Household Longitudinal Study waves 2 to 13 (n = 25,049).** The implication statistic indicates the degree to which a state is indicative of a sequence belonging to the missing or observed group. Dotted lines denote the 95% confidence interval. Missingness was associated with non-caregiving rather than caregiving.

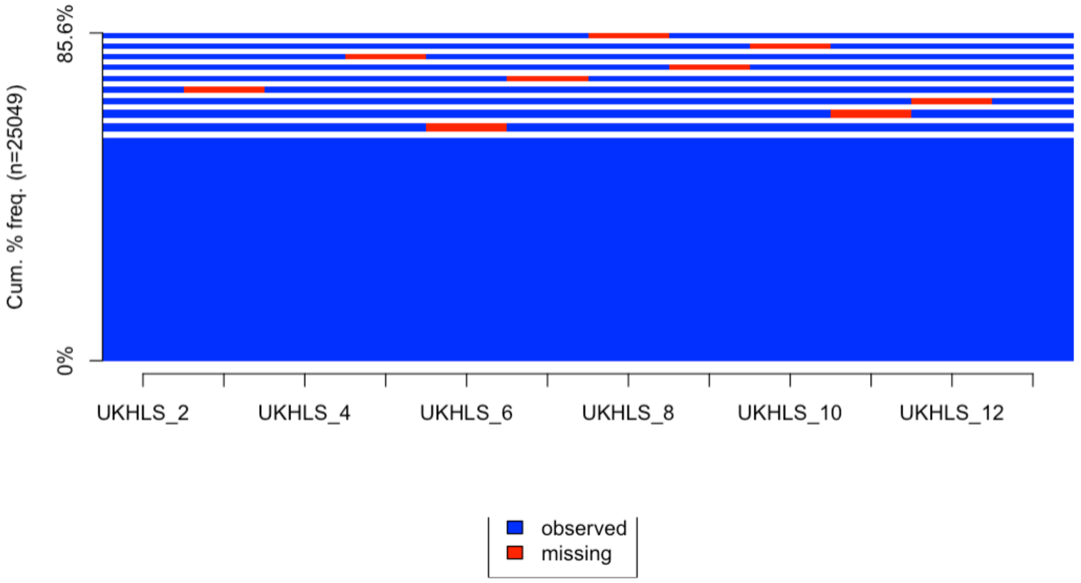

**Figure S. 17: Ten most frequent patterns of missingness in the caregiving status sequence, UK Household Longitudinal Study waves 2 to 13 (n = 25,049).** Observed states are shown in blue and missing states in red. Missingness was sporadic and infrequent, and was not concentrated at the start or end of the observation window.

#### Sequence imputation

Sequence imputation of ten data sets was performed on 25,049 which resulted in a total of 250,490 sequences. Sequence analysis was performed on the aggregated 2,691 distinct sequences.

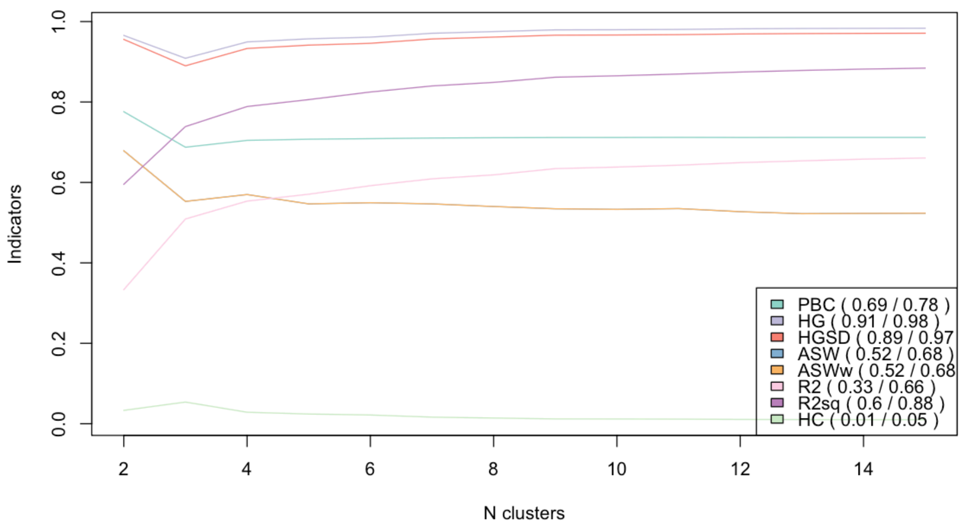

**Figure S.18:** Cluster quality indicators for two- to ten-cluster solutions from optimal matching following sequence imputation (m = 10, n = 25,049). Improvement in fit was gradual across solutions, so the number of clusters was also assessed against the dendrogram and conceptual plausibility.

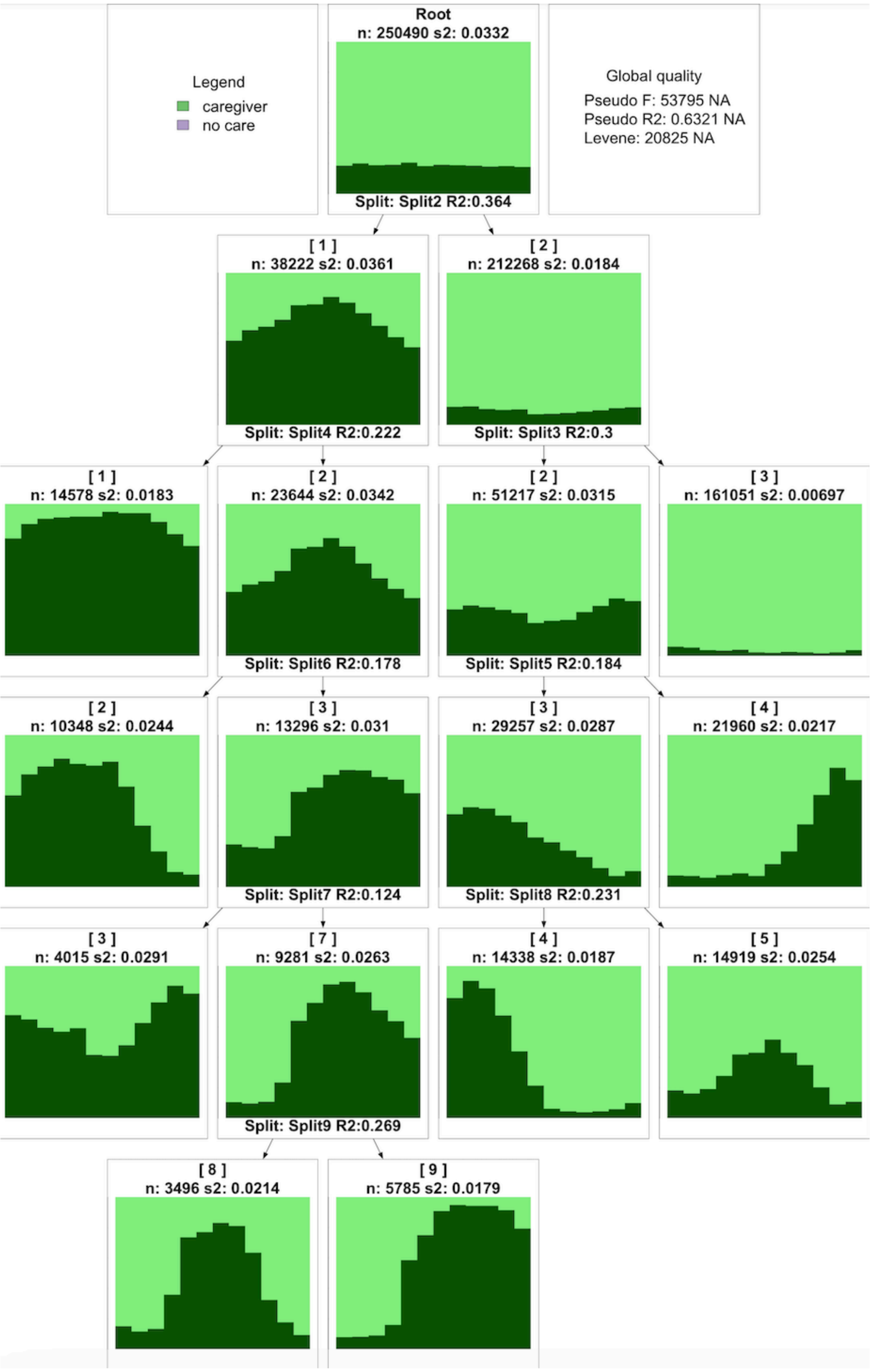

**Figure S. 19:** Cluster tree optimal matching with Ward linkage on the sequence-imputed data (n = 25,049). Eight clusters were retained; splitting the second cluster from the left produced no additional distinct trajectory pattern.

**Description of 8-cluster solution:**

- **Cluster 1: Long-term** caregivers where caregiving is the dominant state throughout all time points.
- **Cluster 2: Former-long** caregivers with long periods of caregiving prior to exit
- **Cluster 3: Recurrent** caregiver with caregiving at start of study, longer break and transition back into caregiving.
- **Cluster 4: Former-short** caregivers with a longer period of non-caregiving after caregiving exit.
- **Cluster 5: Temporary** caregivers, characterised by transition into caregiving and exit.
- **Cluster 6: Emerging-short** caregivers with transition into care and a prior longer episode of non-caregiving followed by a short period of caregiving.
- **Cluster 7: Non-caregivers** with non-caregiving being the dominant state in all waves
- **Cluster 8: Emerging-long** caregivers with transitioning into caregiving followed by a longer period of caregiving.

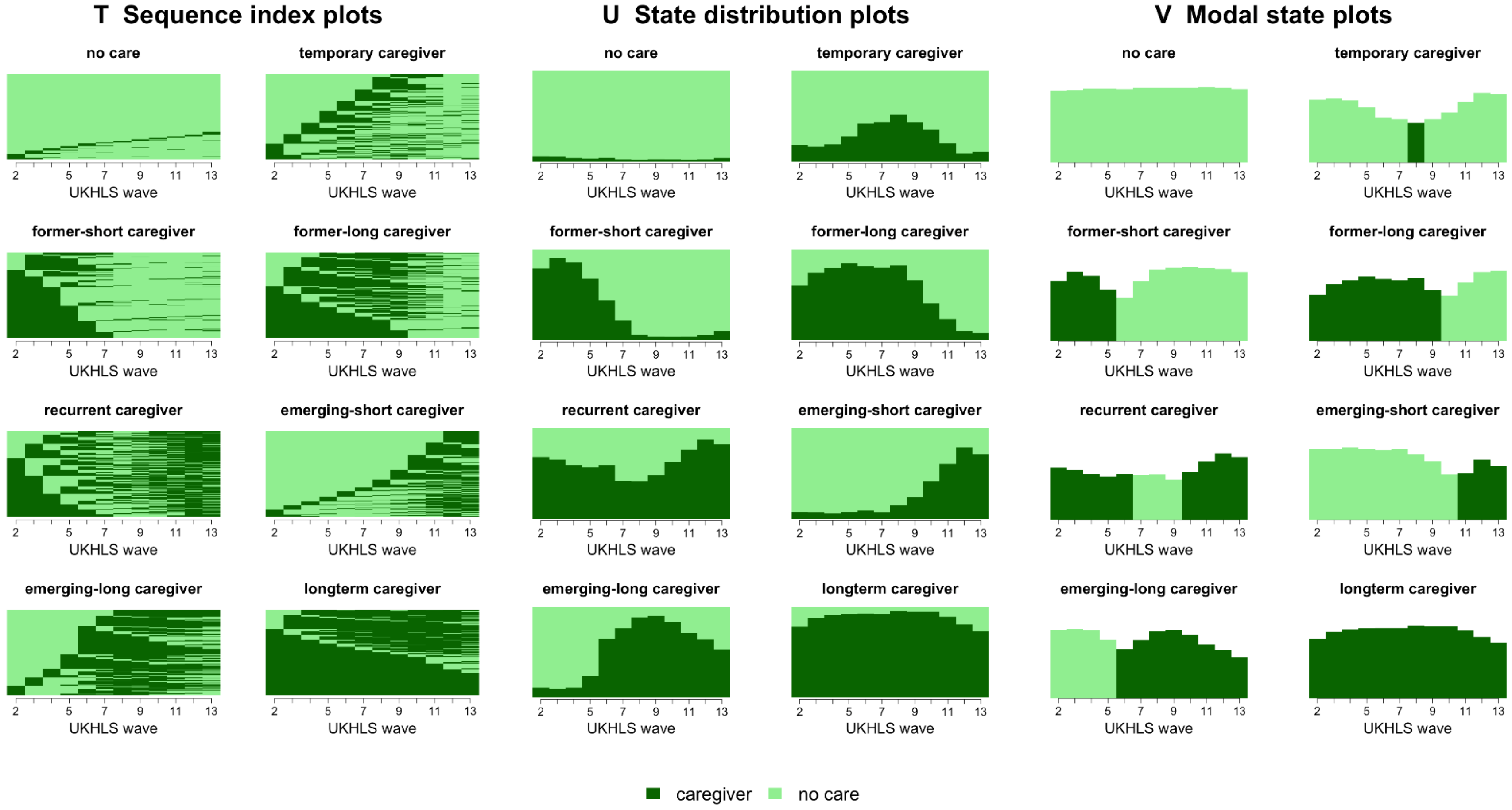

**Figure S. 20:** Caregiving trajectories of the eight-cluster sequence analysis solution, UK Household Longitudinal Study waves 2 to 13 (n = 25,049, m = 10). (T) Sequence index plots, (U) state distribution plots and (V) modal state plots for each cluster derived by optimal matching with Ward linkage on the sequence-imputed data. Dark blue denotes caregiving and light blue non-caregiving. Cluster 3 is not modally in caregiving at the first time point, although panels A and B show that caregiving accounts for a substantial proportion of that cluster at that wave.

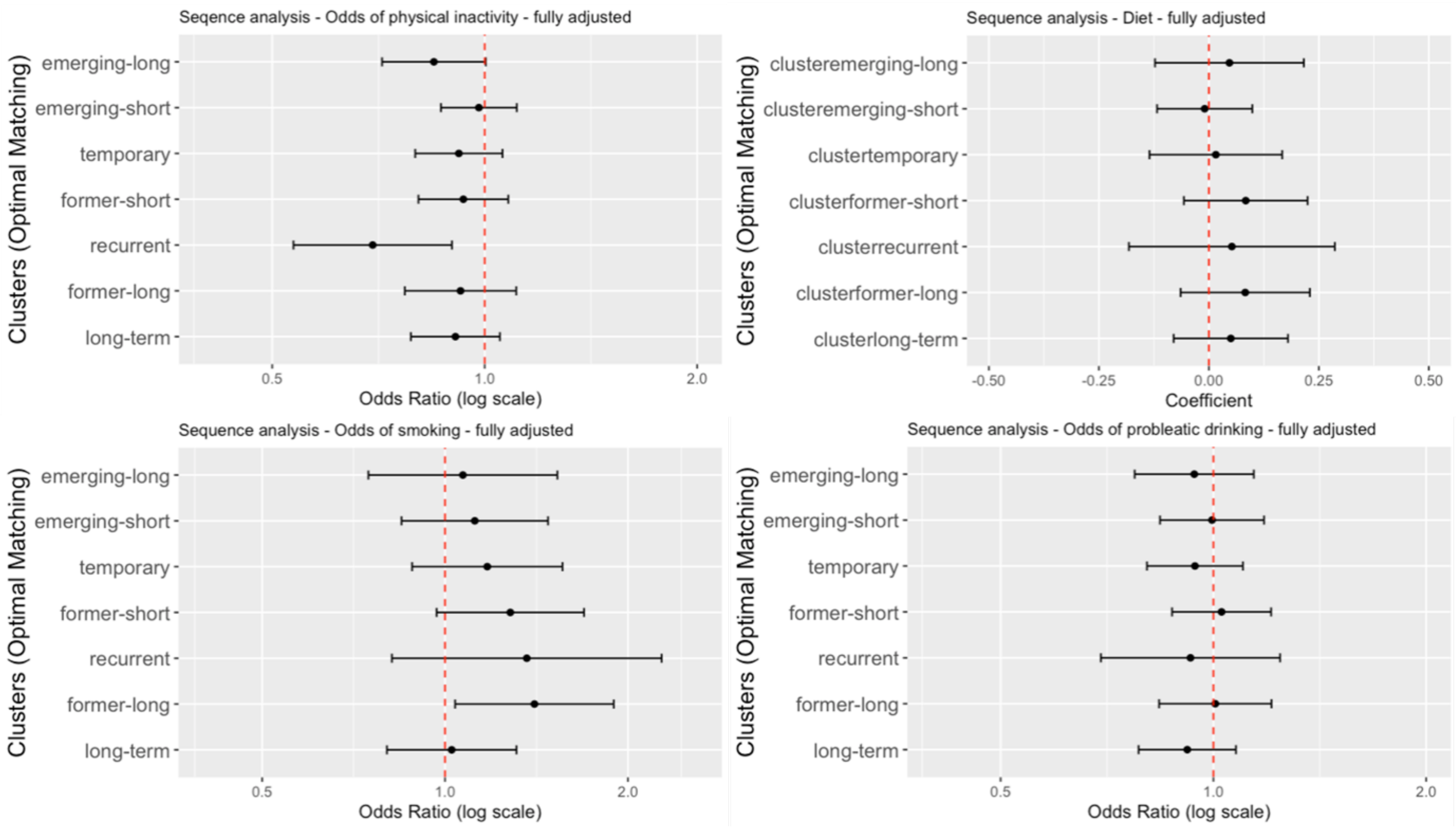

**Figure S.21:** Association between sequence analysis clusters and health behaviours, UK Household Longitudinal Study (n = 25,049, m = 10). Points are odds ratios or linear regression coefficients with 95% confidence intervals from fully adjusted models, pooled across imputations and accounting for the complex survey design and household-level clustering. Non-caregiver is the reference cluster.

### S13: UKHLS Content Plan

**Table S.12:** UKHLS long term plan, adapted from University of Essex

| Module | Waves | | | | | | | | | | | | |
|---|---|---|---|---|---|---|---|---|---|---|---|---|---|
| | **1** | **2** | **3** | **4** | **5** | **6** | **7** | **8** | **9** | **10** | **11** | **12** | **13** |
| **Diet** | | x | | | x | | x | | x | | x | | x |
| **Physical activity** | | x | | | x | | x | | x | | x | (x) | x |
| **Smoking** | | x | | | x | x | x | x | x | x | x | x | x |
| **Alcohol consumption** | | x | | | x | | x | | x | | x | (x) | x |
| **Caregiving** | x | x | x | x | x | x | x | x | x | x | x | x | x |

(x) added mid-field in response to Covid-19 pandemic
